# Multi-omics approach to map a clinically diverse large cohort with homozygous founder RAG1 p.C176F variant among Old Order Mennonites

**DOI:** 10.64898/2026.09.29.26363255

**Authors:** Evan Potts, Marta Toth, Karlla W. Brigatti, Erik G. Puffenberger, Paola R. Suhet, Mustafa Talib, Alanna E. Koehler, Zsuzsanna Gaál, Kellie A. Larsen, Rahim Z. Miller, Daryl J. Gibson, Philip Mendez, Zixiao An, Jeffrey J. Bednarski, Peter Blazso, Marita Bosticardo, Sharat Chandra, Ivan K. Chinn, Joelle W. Clark, Antonio Condino-Neto, Ottavia M. Delmonte, Dimana Dimitrova, Emily S. J. Edwards, Ganesh V. Halade, Jennifer R. Heimall, Ayal Hendel, Sarah E. Henrickson, Katherine E. Herman, Avni Joshi, Heather Kenney, Maleewan Kitcharoensakkul, Gloria Magro, Eliza C. Martin, Edgar B. Oliveira-Junior, Mei-Sing Ong, Oscar E. Ospina, Francesca Pala, Kady Palmer, Jason P. Raasch, Mathilde Salamon, Christopher D. Scharer, Christine M. Seroogy, Svetlana O. Sharapova, Raz Somech, Michael Teng, Timothy J. Thauland, Boglarka Ujhazi, James W. Verbsky, Brant R. Ward, Shengfeng Xu, Melis Yilmaz, Yuhang Zhang, Wei Zhao, Krisztian Csomos, Yu Nee Lee, David G. Schatz, Manish Butte, Nicholas L. Rider, Laura E. Poskitt, Luigi D. Notarangelo, Kevin A. Strauss, Jolan E. Walter

## Abstract

Partially preserved function of recombination-activating genes (RAG) 1 and 2 can be linked to divergent and hard-to-predict clinical and immunological phenotypes, even among close relatives. It is unclear how intrinsic RAG hypomorphism interacts with extrinsic environmental pressures to shape clinical trajectory. We dissect this phenomenon in a unique cohort of 18 Old Order Mennonite individuals with a novel missense founder *RAG1* variant (p.C176F) with 25.8% recombinase activity of wild-type protein. After studying a myriad of markers, we demonstrate varying severity and progression of clinical disease (predominantly in females) that paralleled decline in CMV-specific immune competence, break in tolerance with hallmark presence of antibodies to IFNα, a TH1/IFNγ-driven inflammatory milieu with elevated serum markers (CXCL9/10), expansion of follicular helper T cells and clonal B cells enriched for autoreactive-prone VH4.34 B cell receptor. BCR and TCR repertoires carry the fingerprints of both extrinsic and RAG-driven intrinsic changes. Through this ‘experiment of nature’, our cohort demonstrated that partial RAG deficiency (pRD) is not a monolithic disease but the result of compounded RAG intrinsic and extrinsic events. Overall, this study uses multi-omics approaches to dissect the contribution of a hypomorphic RAG variant to variable severity of clinical disease and immunological dysregulation at cellular and molecular level. Emerging biomarkers of a multi-omic roadmap should allow monitoring of progression of clinical disease and guide who and when to treat with hematopoietic stem cell transplant in pRD.

**One sentence summary:** A unique cohort of pediatric and adult US Mennonite patients with shared novel partial RAG defect and variable penetrance of disease allow correlation of clinical severity with RAG-intrinsic and extrinsic factors driving plasticity in T and B cell competence to eliminate infections and tolerance to self.

## INTRODUCTION

The recombination-activating genes 1 and 2 (*RAG1/2*) encode lymphoid-specific proteins primarily expressed in T and B cell precursors to orchestrate the generation of diverse primary T and B cell receptor (TCR/BCR) repertoires and to foster elimination of polyreactive clones at central and peripheral tolerance checkpoints (*1–4*). Biallelic pathogenic *RAG1/2* variants impede V(D)J recombination to varying extents and are associated with a broad spectrum of clinical and immunological phenotypes in humans. The clinical landscape includes patients spanning all ages from asymptomatic stage to other who develop infections and/or immune dysregulation, including autoimmunity, chronic inflammation, lymphoproliferation and even malignancy (*5–7*). In parallel, immunologic phenotypes span a wide spectrum; from full absence of T and B cells (severe combined immunodeficiency [SCID]) to ones with inflammatory state such as T^+^B^-^ Omenn syndrome (OS) and T^low^B^low^ combined immunodeficiency with granuloma and autoimmunity (CID-G/AI) to cases with mild T cell lymphopenia and antibody deficiencies of varying severity (*5–7*). The primary driver of the variable clinical and immunological landscape of RAG deficiency has been attributed to the amount of residual RAG protein activity, with a broad genotype-phenotype correlation established (*8*). Patients with residual recombinase activity are unified under the term of partial RAG deficiency (pRD). Accumulating evidence suggests that genetic modifiers and environmental factors (e.g. viral infections, dysbiosis) substantially shape immune dysregulation (*4, 9–11*). In particular, viral infections are likely a major player in triggering inflammation, autoimmunity and clonal expansion in many variants of RAG deficiency, such as OS, TCRγδ leaky-SCID (γδLS) and CID-G/AI phenotypes (*12–14*). Human herpesviruses such as Cytomegalovirus (CMV), Epstein-Barr virus (EBV) and Varicella Zoster virus (VZV), have been reported to shift clinical and immune phenotype in pRD patients, including clonal expansion and misdirected immune responses with a broad spectrum of autoantibodies, including those targeting cytokines (*13, 15*). Mechanistically, we proposed that RAG-driven stochastic primary BCR/TCR repertoire changes in pRD might induce decreased competence to react against infectious agents with increased antigen load that might lower tolerance filters and contribute to the progression of peripheral immune dysregulation (*4*). Antigen-specific adaptive immune responses and declining competence against infectious agents has, however, been less investigated. Assessment of individuals for progression of loss of competence and tolerance from the same family sharing the same *RAG* genotype could provide clearer insight into the palette of extrinsic factors shaping the variable clinical and immunological expressivity (*7, 9*).

The introduction of Newborn Screening for SCID (SCID-NBS) in the United States enables early diagnosis of some variants of RAG deficiency and considered to be the main driver to improving overall survival for SCID patients (*16*). The test screens for severe T cell lymphopenia and it is built on the absence of T cell receptor excision circles (TRECs), a by-product of V(D)J recombination in developing thymic T cells (*17, 18*). SCID and OS infants with RAG variants of near absent recombinase activity and TRECs are readily detected through positive SCID-NBS and can be fast-tracked to hematopoietic stem cell transplant (HSCT) (*19*). However, it is yet unknown whether all pRD patients can be identified by SCID-NBS given the variability of T cell lymphopenia. It has been described that a subset of pRD patients receives HSCT later in life (*20, 21*). As the clinical trajectory of these pRD patients is heterogenous with unclear prognosis, there is unmet need for biomarkers to better understand timing of definitive therapy (*20*, *21*).

To model this conundrum, we share our studies of the largest human pRD cohort from the Old Order Conservative Swiss Mennonites (OOCSM) carrying the same novel variant (RAG1 p.C176F) with variable clinical and immunological expressivity to translate the clinical severity to the cellular level utilizing multi-omics approaches including proteomics, T and B cell repertoire analyses and CMV-specific T cell functional assays. We aim to elucidate the palette of RAG-intrinsic and extrinsic influences shaping this phenomenon in a highly vulnerable community where access to definitive therapy is limited.

## RESULTS

### RAG1 c.527G>T p.C176F is a pathogenic founder variant among OOCSM

Through a multi-institutional collaboration, we identified 18 patients homozygous for the same *RAG1* variant (c.527G>T, p.C176F). These patients originate from geographically distinct OOCSM communities across nine different U.S. states and share a rural, agrarian lifestyle (*22*).

To determine whether this variant is enriched in OOCSM populations, we analyzed whole-exome sequencing data from 828 randomly selected OOCSM adults from southeastern Pennsylvania. Forty-nine individuals were heterozygous (Hets) carriers, corresponding to a minor allele frequency (MAF) of 2.96 x 10^-2^, approximately 10^5^-fold higher than the MAF in the non-Finnish European population (GnomAD v4.1.0, 4.24 × 10⁻⁶; *p* < 0.0001) (**Fig1A**). We constructed a multi-generational pedigree, which demonstrated segregation of this variant across multiple ancestral lineages within the OOCSM population (data available upon request). This extended pedigree includes individuals from several distinct yet genetically related Mennonite demes (i.e., the Groffdale, Weaverland, Reidenbach, and Hagerstown Mennonite groups). Despite geographic and generational separation, these lineages converge on a common ancestral origin.

**Figure 1.**
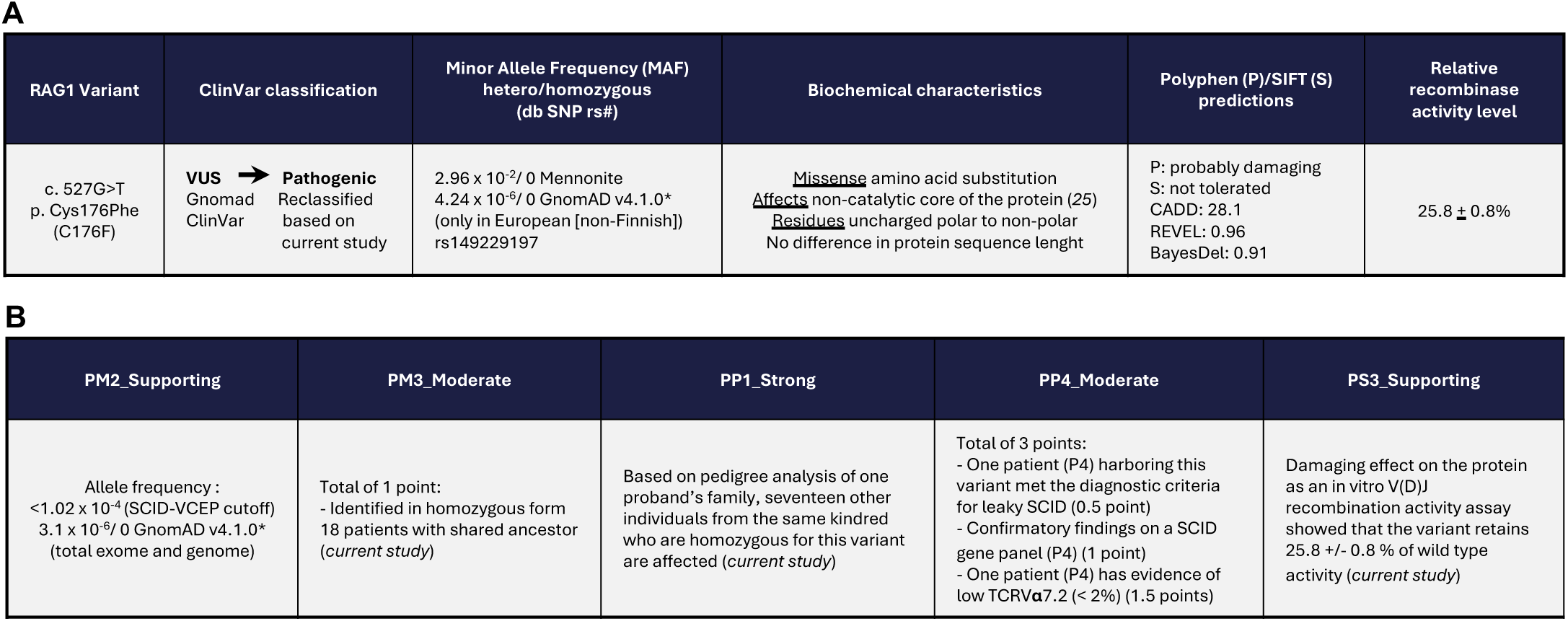
Segregation of RAG1 p.C176F across Mennonite families in the United States. Segregation of the homozygous RAG1 c.527G>T (p.C176F) variant among 18 affected individuals with partial RAG deficiency (pRD) from multiple Mennonite families across the United States. A) Mapped geographic distribution. B) Shared ancestry of affected patients included in this cohort. C) Characteristics of novel RAG1 variant that is being reclassified from VUS to pathogenic. (*based on gnomAD 03/2026). D) Variant resolution based on variant curation expert panel (VCEP) specifications version 2.1.0.

Previous case reports of the *RAG1* p.C176F missense variant has not fully established pathogenicity (*15*, *23*). Several *in silico* predictive tools consistently suggest this variant as probably damaging (*24*) (**Fig1A**).

Notably, a recent phylogenetic and structural analysis demonstrates that cysteine 176 is a highly conserved zinc-coordinating residue in the first position of a CxxC motif in a previously unrecognized N-terminal zinc-binding domain (NZD) of RAG1 (*25*). Mutation of cysteine to phenylalanine is expected to disrupt zinc coordination and to compromise NZD structure and function (*25*)and mutation of other NZD zinc-coordinating residues disrupts NZD structural stability and significantly reduces recombination signal sequence (RSS) mediated V(D)J recombination (*25*). Misfolding of NZD might indirectly affect the stability of the RAG1 protein and/or the function of other domains of RAG1 including the catalytic domain.

Relative recombinase activity was previously unpublished for this variant and so was evaluated using a previously described platform in Abelson virus-transformed pro-B-cell lines expressing the RAG1 variant (*8*). *In vitro* functional assessment of the p.C176F variant showed a residual recombinase activity of 25.8 +/- 0.8 % of wild type activity, consistent with a hypomorphic effect (assay done in triplicates, Notarangelo lab, see in Materials and Methods below).

Initially, the Variant Curation Expert Panel (VCEP) for SCID (supported by ClinGen) assessed the NM_000448.3:c.527G>T p.Cys176Phe as variant of uncertain significance (VUS) in 2024-01-17, version 1.0 and 1.1. Based on our current cohort, the new ACMG/AMP and ClinGen SCID VCEP assessment classified the variant pathogenic as based on cumulative assessment of (1) PM2_Supporting, (2) PM3_Moderate, (3) PP1_Strong, (4) PP4_Moderate, (5) PS3_Supporting (VCEP specifications version 2.1.0) (**Fig1B**).

Collectively, with all these variant curation efforts, functional and structural studies support that RAG1 p.C176F is a pathogenic founder variant in the OOCSM population, likely introduced by a single Mennonite immigrant to Pennsylvania in the 18^th^ century. Consequently, this *RAG1* variant was incorporated into the PlainInSight genetic screening panel (Clinic for Special Children in Pennsylvania, March 2025) for targeted sequencing to facilitate the detection of both heterozygous carriers and homozygous cases within the communities of Old Order Mennonites in southeastern Pennsylvania and beyond.

### Improved molecular diagnosis and definitive therapy for pRD with implementation of SCID Newborn Screening (SCID-NBS)

Our cohort of 18 patients (P1-P15, cP1-cP3) was divided into those who were born after and prior to the implementation of SCID-NBS in their state.

Ten patients (P6-15) (5 males and 5 females; pre-SCID-NBS cohort) were born before the implementation of SCID-NBS including the first published case diagnosed in their 10’s (*15*). Five patients (4 males and 1 female) were asymptomatic or mildly symptomatic at the time of the diagnosis. Three females received HSCT (**Table 1**).

Eight patients (P1-5,cP1-cP3) (3 males, 5 females) had a positive SCID-NBS as the trigger for diagnosis in their home state, including Pennsylvania (n=3), Wisconsin (n=2), New York (n=2), and Missouri (n=1). TRECs (count/μl) were variable, ranging from no amplification to just below the cutoff. Five of eight patients received HSCT (**Table 1**).

In comparison to the pre-SCID-NBS group, children diagnosed in the era SCID-NBS had molecular diagnosis in a significantly younger age (1.4 vs 23.3 years, p=0.0001), which contributed to performing HSCT earlier (mean 3.1 vs 25.3 years, p=0.04) and in a higher proportion of patients (62.5% vs 30%), with fewer pre-transplant co-morbidities. Nevertheless, early CMV infection remains a potentially fatal complication. As of 2026, 12 (80%) of 15 patients are alive. Three patients died, including two post HSCT (**Table S1**).

### Longitudinal clinical outcome varies in severity by age and sex in pRD cohort

We characterized the severity of clinical course in 15 patients (P1-P15) at multiple timepoints longitudinally (**Fig2A**), excluding three individuals (cP1-cP3) who had significant confounding genetic comorbidities (**Table S1**).

**Figure 2.**
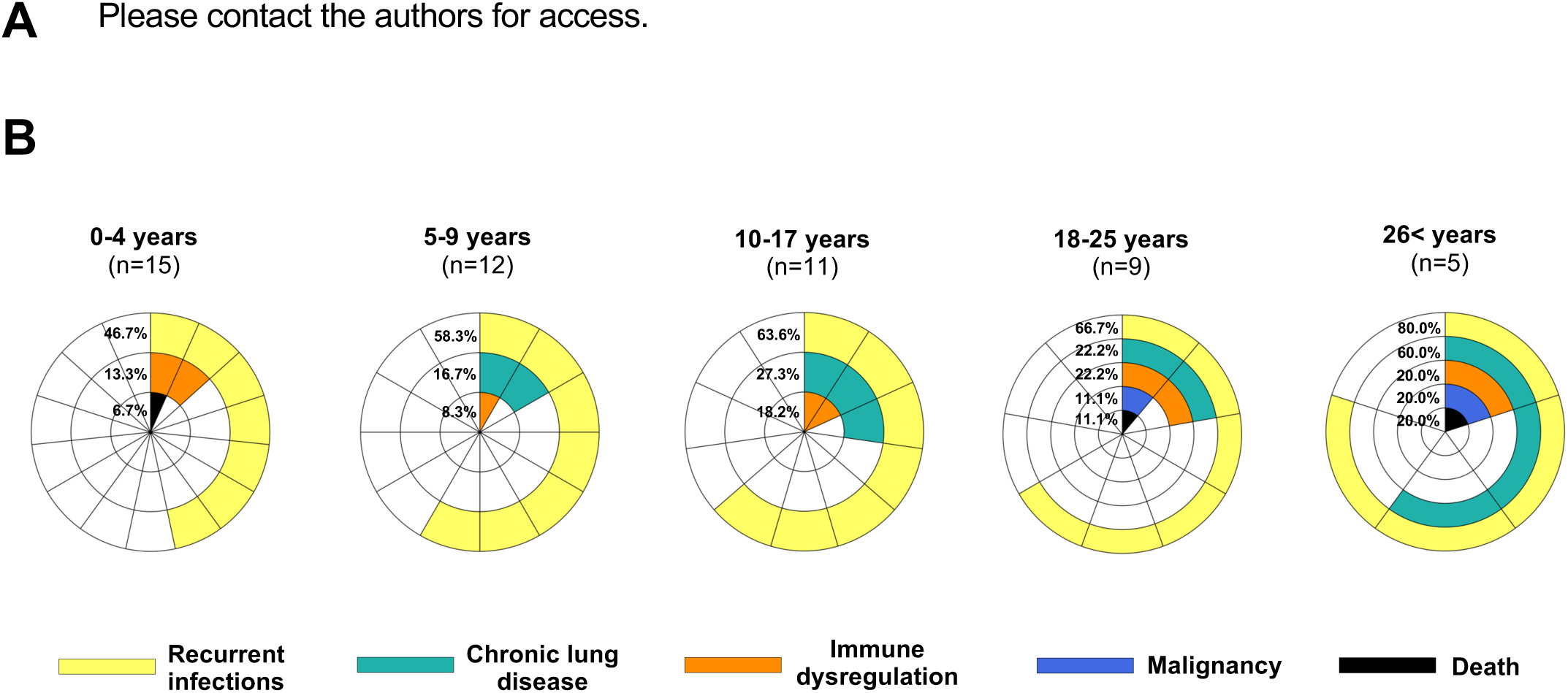
Major clinical manifestations and comorbidities in pRD patients by age. **A)** Swimmer plot depicting timing of clinically relevant events, hematopoietic stem cell transplantation (HSCT) in patients grouped by the era of pre- or post-initiation of newborn screening for severe combined immunodeficiencies (SCID-NBS). **B)** Clinical manifestations of pRD across five age groups. Frequency of infections and the prevalence of immune dysregulation and organ damage increased with age. Immune dysregulation category included autoimmune manifestations, inflammation and granulomas.

Initial clinical manifestations were recurrent infections (bacterial, fungal, parasitic and viral causes, most notably CMV [n=3] and VZV [n=4] **(Fig2B)**(**Table S1**). With age, non-infectious manifestations accumulated, including chronic lung disease (n=5, 9-28 years of age), immune dysregulation (n=6, 1.2-26 years of age) and malignancy (n=1, age 20’s). Above 26 years of age the prevalence of chronic lung disease reached 60%. Other common features of immune dysregulation included inflammatory bowel disease (IBD) and granulomas (lung, skin and esophagus).

In order to distinguish clinically relevant subgroups of patients based on reproducible and objective measures, Immune Deficiency and Dysregulation Activity score v.2.2 (IDDA) was applied at each time point of clinical assessment (*26*, *27*). This scoring system that was previously validated in Cytotoxic T-Lymphocyte-Associated protein 4 (CTLA4) and Lipopolysaccharide Responsive Beige-like Anchor protein (LRBA) deficiencies which disorders also present with variable clinical penetrance of infectious and non-infectious complications (*26*, *27*). A total of 73 timepoints from 15 patients were scored, including 65 from the pre-HSCT period. K-means clustering of the pre-HSCT timepoints stratified clinical severity into three clusters corresponding to mild (IDDA<9.41, n=25), intermediate (IDDA 9.41–34.75, n=22), and severe (IDDA≥34.75, n=18) disease **(Fig3A)**. When the highest IDDA score timepoints per individual were selected, the mean age was the lowest in the mild (13.2 years) versus moderate (21.2 years) and severe (16.8 years) clusters. These severe cluster consisted predominantly of females (85.7%), whereas the moderate cluster included only males (n=4) suggesting a sex-biased disease progression pattern (p=0.03)(**Fig3B**). We further assessed the relationship between age, sex and disease severity (IDDA scores) (**Fig3C**). In the full dataset, linear regression revealed significant opposite age-related trends in females vs males (β = −1.125, p = 0.02)**(Fig3C left panel)**. After excluding P1 and P2 with severe CMV disease the difference remained significant for positive correlation with age in females vs males (ρ = 0.58, p = 0.002)(**Fig3C right panel**).

**Figure 3.**
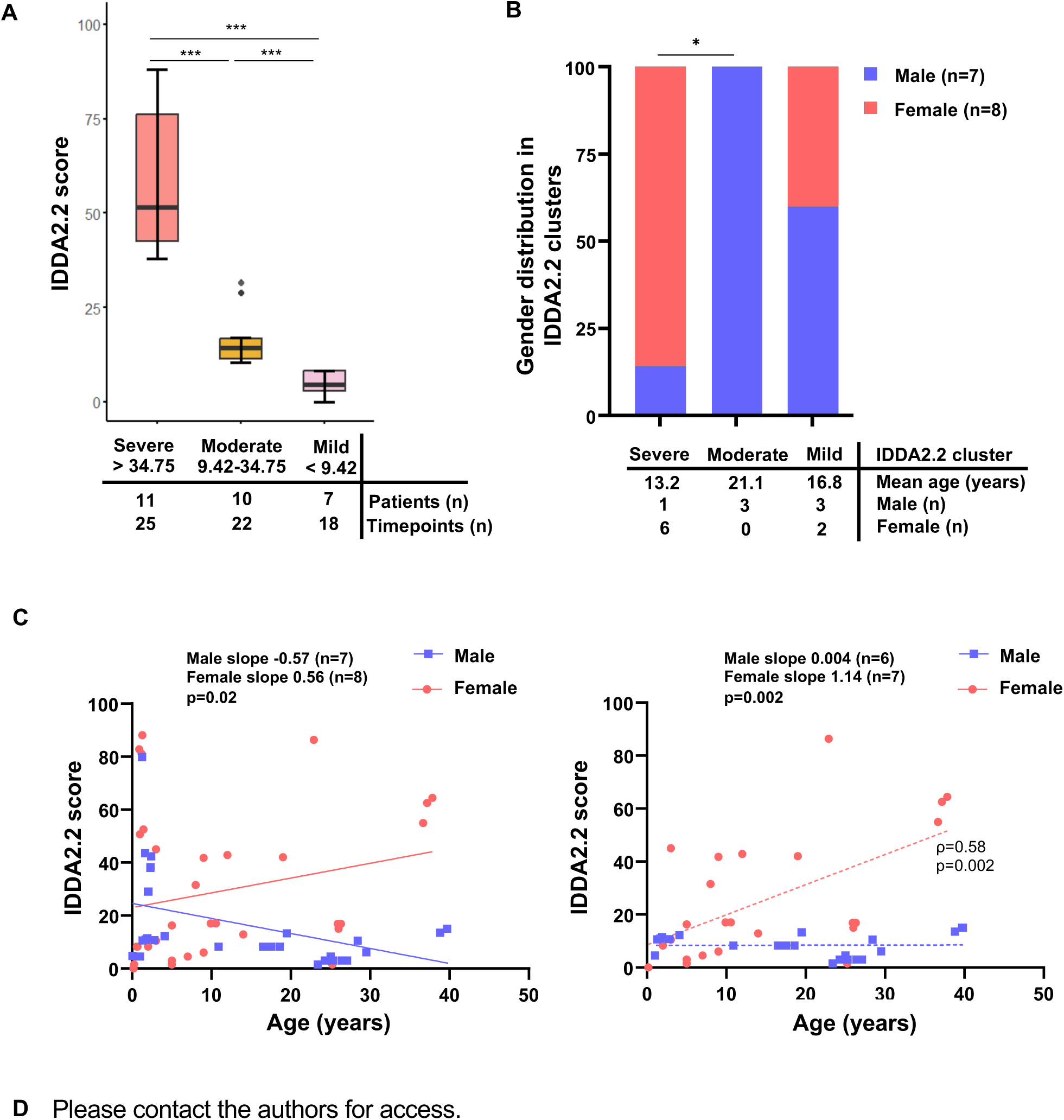
Evaluation of clinical severity of immune dysregulation at different time points in pRD patients. **(A)** K-means clustering of Immune Deficiency and Dysregulation Activity (IDDA) 2.2 scores, including each time point of clinical assessment. **(B)** To avoid sampling bias, we assigned the highest IDDA2.2 score to each patient and categorized them by severity. **C)** Pre-HSCT IDDA2.2 scores (from all available time points per patient) were plotted against age and stratified by sex. Slope analyses were performed both including patients with severe early CMV infection (P1, P2) (**left panel**) and after their exclusion (**right panel**). **D)** IDDA2.2 scores of the peri-HSCT period showed a decreasing trend during follow-up (peri-transplant vs. most recent follow-up scores per patient), except for the two patients who died (P1, P6). Slopes were analyzed by linear regression; correlation was calculated by Spearman rho test.

Supportive therapy was not uniform: only a fraction of patients received prophylactic antibiotics (40%), immunoglobulin replacement therapy (IgRT) (46.7%) or immunomodulatory treatment (46.7%)] (**Table S1**).

At the time of HSCT, IDDA scores were highly variable (mean 25.84, range 0-64.4) with most patients in the severe IDDA cluster (p=0.03)(**Fig3D**). Among those who survived HSCT, IDDA scores showed a decreasing trend during follow-up (mean 6.15, 0-17.2) in a period of average of 3.9 years (range 1.44-8 years). Two patients (P1, P6) died post-HSCT and marked increases in IDDA scores (>100) reflected clinical deterioration. Patients with mild to moderate disease severity, fared well post-HSCT, demonstrating substantial clinical improvement. P2 is the only survivor with a high pre-HSCT IDDA score of 50 (**Fig3D**).

### IFNγ-driven soluble inflammatory factors correlate with clinical severity in pRD

It has been previously described that distinct RAG phenotypes share a common inflammatory signature (*11*), however in practice, patients demonstrate variable clinical features (*3*). We aimed to associate our clinical grading system (IDDA clusters) with 20 soluble factor levels including those previously described in pRD (*11*). Using Legendplex flow-based method, we confirmed in pRD a pattern primarily dominated by Th1-type cytokine and chemokines (IFNγ, CXCL9, IP-10/CXCL10) (**Fig4A**). Th2 cell-recruiting and polarizing factors (CCL17, CCL22, IL-13) were also elevated in pRD vs Hets and HDs which may reflect a mixed T-cell response (**Fig4A)**. Among top plasma mediators only IP-10/CXCL10 **(Fig4B-C)** and CXCL9 **(Fig4D-E)** levels correlated positively with the IDDA scores in pRD (**FigS1A)**. Elevated CXCL9 and CXCL10 were also confirmed by ELISA and trended with IDDA score, however significance was only reached for CXCL9 **(FigS1B-C)**. In our cohort, plasma IFNγ concentration was near the detection limit both with LegendPlex assay and ELISA **(FigS1D)**.

**Figure 4.**
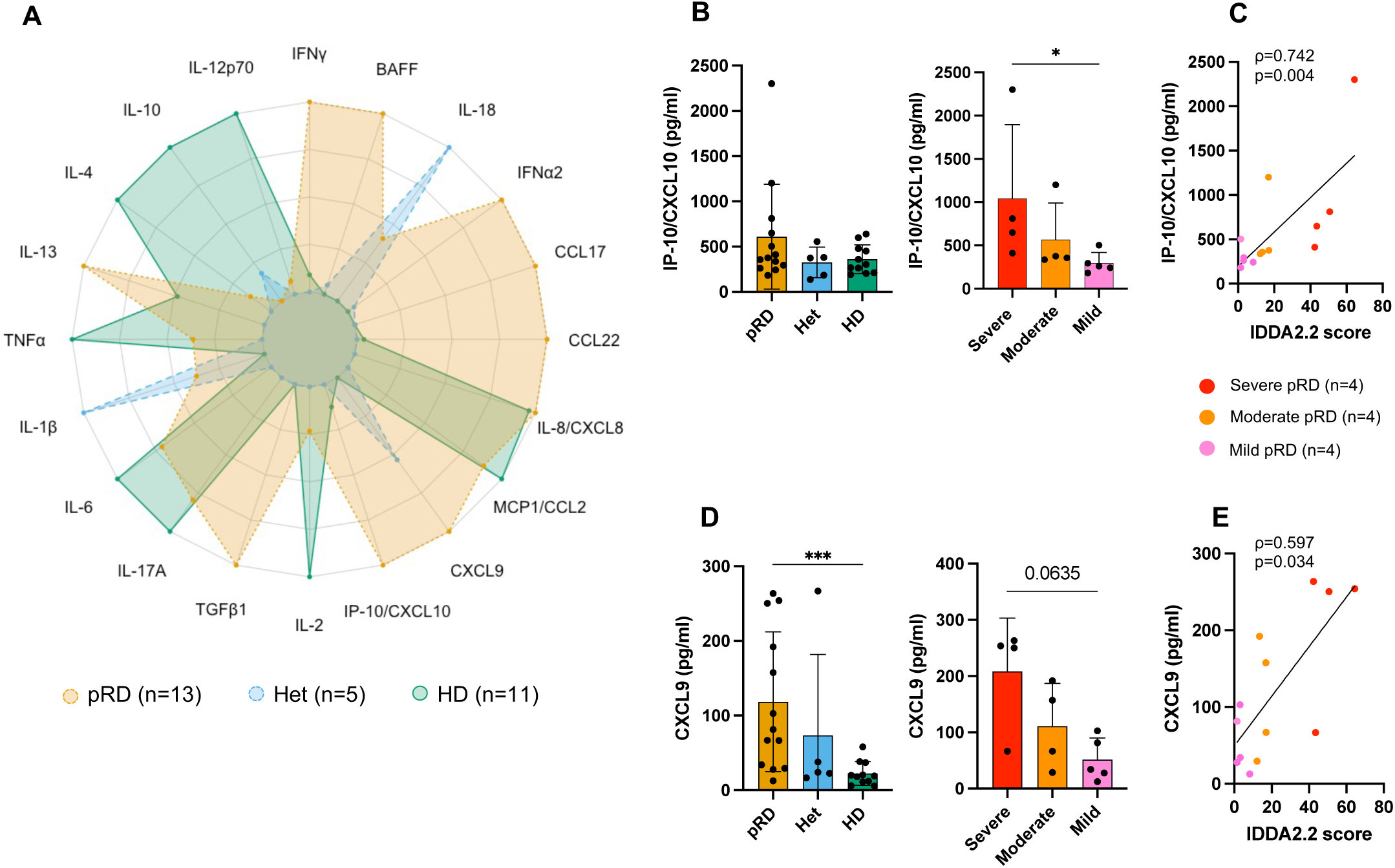
IFNγ pathway-driven inflammatory response correlates with clinical severity in pRD patients. **(A)**. Spider plot diagram of 20 plasma factors measured from samples obtained 13 timepoints from 11 pRD patients; 5 heterozygous carriers (Hets) and 11 HDs. Values are scaled to the maximum and minimum value of each biomarker. **B)** Comparison of IP-10/CXCL10 level between **(left panel)** pRD, RAG1 carriers and HDs; (**right panel)** severe, moderate and mild pRD patients. **C)** Correlation between IDDA2.2 scores and plasma IP-10/CXCL10 levels. **D)** Comparison of CXCL10 level between **(left panel)** pRD, RAG1 carriers and HDs; **(right panel)** severe, moderate and mild pRD patients. **E)** Correlation between IDDA2.2 scores and plasma CXCL9 levels. Significance was calculated using Mann-Whitney tests; p≥ 0.05 was considered statistically significant. Correlation and p value was calculated by Spearman correlation and *p* test indicated by the ρ correlation coefficient. P8 was excluded from the BAFF level analysis due to Rituximab treatment. Abbreviations: IP-10: interferon gamma-induced protein 10, MCP1: monocyte chemoattractant protein 1, TGFβ1: transforming growth factor beta 1, TNFα: tumor necrosis factor alpha.

Further, we tested 58 lipid mediators known to contribute to the inflammatory and inflammation-resolving processes **(FigS1E)**. We performed sparse Partial Least Squares Discriminant Analysis (sPLS-DA) to discriminate the pRD, Het and HD groups **(FigS1F)**. Among the top 10 lipid mediators contributing to the Component 1 **(FigS1G)**, only angiogenic 11,12 Epoxyeicosatrienoic acid (11,12 EET) correlated positively with the IDDA scores (p=0.015)**(FigS1H)**.

These findings confirmed our previous observation that pRD patients display a common dysregulated T_H1 I_FNγ-driven inflammatory signature (*11*). Furthermore, we demonstrated a positive correlation of IFNγ-driven CXCL9/CXCL10 level with worsening clinical state, providing potential biomarkers for disease progression.

### Dynamics of B cell receptor (BCR) repertoire and immune subset composition with clinical disease severity in pRD

To correlate the B cell dysregulation with the clinical severity, we performed peripheral B cell-focused multiomics approaches including in-depth immunophenotyping (**FigS2A**), bulk IGH sequencing for BCR repertoire assessment and measuring the level of plasma neutralizing IFNα antibodies. B cell count was preserved among young patients (<10 years of age, 5 timepoints) while older patients displayed progressive B cell lymphopenia with a decreasing frequency of CD38^hi^CD24^hi^ transitional B cells (TrB) regardless of clinical state (>10 years of age, 8 timepoints) (**Fig5A-C**). We noted a significantly lower fraction of TrB cells when compared between adult pRD patients and HDs (**FigS2B**), consistent with our previous work (*4*).

BCR repertoire diversity (visualized by tree maps), Shannon and D50 indices were comparable between pRD patients, Hets and HDs **(Fig5D-F)**. However, with increased age, BCR repertoire diversity decreased **(Fig5E-F)**, whereas rate of somatic hypermutation (SHM) **(Fig5G)** and frequency of top 10 clones **(Fig5H)** were increased as seen in adult vs pediatric pRD patients. Adult pRD patients also had elevated level of activated “naïve” B cells over time **(Fig5I, FigS2E)**. In our prior work, activated “naïve” B cells accumulated SHM (*4*), and therefore likely contribute to the SHM and oligoclonal expansion in our current cohort. Neither indices of diversity, SHM **(FigS2F)** nor clonality rates **(FigS2G)** in total BCR repertoire correlated with IDDA scores.

**Figure 5.**
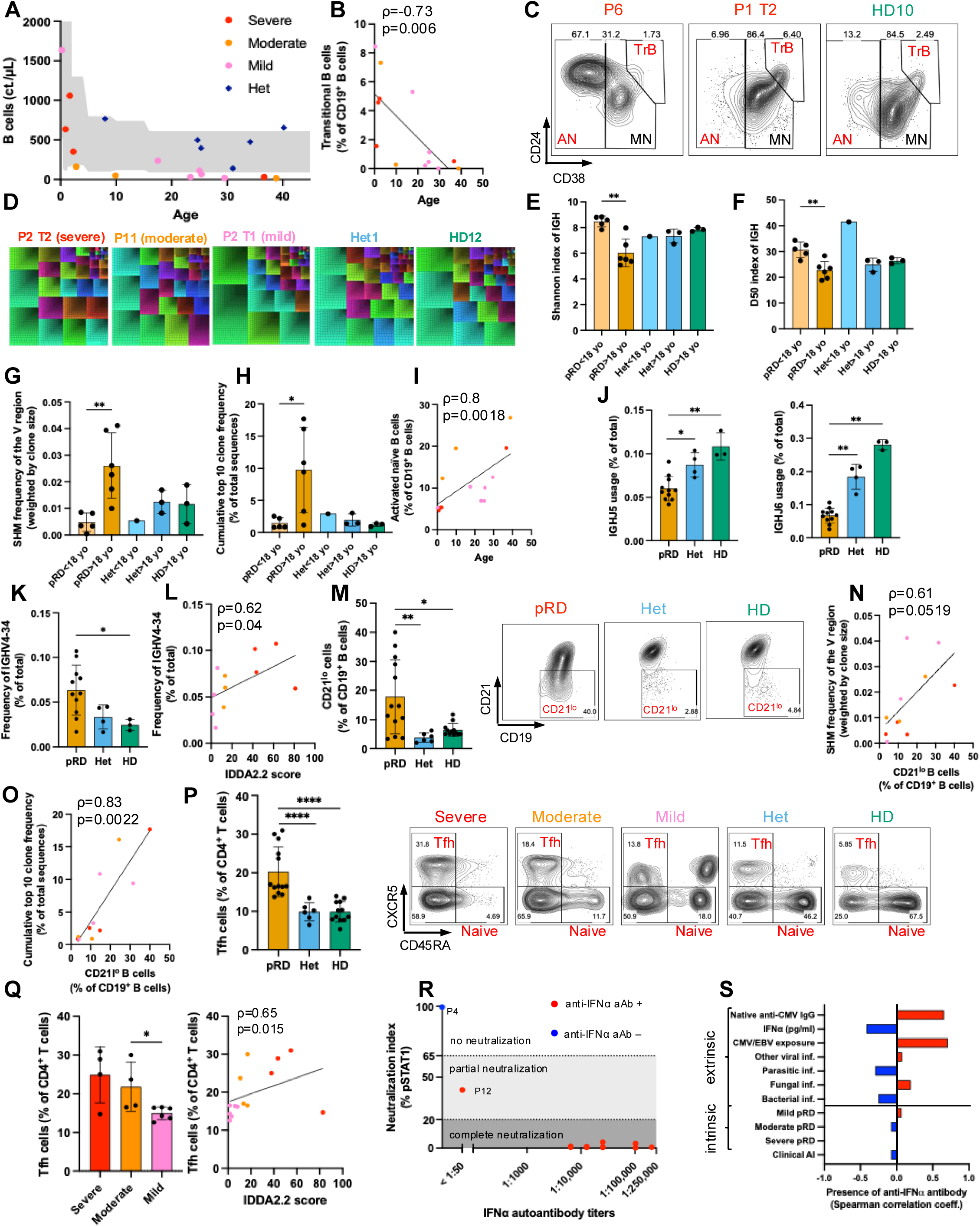
BCR repertoire disturbance shows correlation with disease severity in pRD. Analysis includes 12 timepoints of 10 pRD patients. **A)** B cell counts. **(B)** Correlation between TrB and age. **C)** Flow plots for TrB, MN and AN B cells. **D)** Treemaps for IGHV gene frequencies (top 2500 clones). **E)** Shannon index. **F)** D50 index. **G)** SHM rate. **H)** Top 10 clones. **(I)** Correlation between AN B cells and age **J)** Frequency of IGHJ5 and IGHJ6. **K)** Frequency of IGHV4-34. **L)** Correlation between IGHV4-34 and IDDA2.2 scores. **M)** CD21^lo^ B cell frequency with flow plots. **N)** Correlation between SHM rate and CD21^lo^ B cells. **O)** Correlation between top 10 clones and CD21^lo^ B cells. **P)** Tfh frequency with flow plots. **Q)** Frequency of Tfhs and correlation with IDDA2.2 scores**. R)** IFNα neutralization. **S)** Correlation between anti-IFNα antibody and extrinsic and intrinsic factors. P≥0.05 was considered statistically significant by Mann-Whitney test. Correlation and p value was calculated by Spearman *ρ* and *p* test. Abbreviations: AN: Activated Naïve; MN: Mature Naïve; Trans: Transitional; Tfh: Follicular Helper T; SHM: somatic hypermutation; pSTAT1: phosphorylated Signal Transducer and Activator of Transcription 1. One patient was excluded because the raw input sequences were too low (n=68).

As a shared intrinsic feature in RAG deficiency, the distal J gene segments, IGHVJ5 and J6 were underrepresented in BCR repertoire pf all pRD patients **(Fig5J, FigS2H)** vs Hets and HDs as we previously described (*4*, *28*). In contrast, usage of *IGHV4-34* was significantly higher in pRD patients vs HDs **(Fig5K, FigS2H)** and also correlated with IDDA scores **(Fig5L)**. The preferred use of this gene segment has not been described in relation to RAG-driven effect, but published pRD cases on protein (9G4 antibody) or gene segment level (*IGHV4-34* BCR)(*4*, *11*, *29*). As a marker of immune dysregulation, *IGHV4-34* has been widely seen in autoimmune diseases and known to bind to an erythrocyte antigen (*30*) and various antigens including ssDNA and dsDNA (*31*).

Among B cell markers of immune dysregulation, we observed significant expansion of CD21^lo^ B cells without correlation with clinical severity in pRD patients vs Hets and HDs **(Fig5M, FigS2I)**. Furthermore, frequency of IGH SHM **(Fig5N),** usage of *IGHV4-34* (**(FigS2J)** and the top 10 clones **(Fig5O)** correlated positively with the expansion of CD21^lo^ B cells. These findings parallel Tfh expansion in pRD **(Fig5P)**, however, unlike for CD21^lo^ B cells, Tfh expansion correlated with clinical severity **(Fig5Q)**. This phenomenon likely reflects aberrant Tfh-CD21^lo^ B cell crosstalk in extrafollicular niche and may promote dysregulation (*32*, *33*).

As a marker of autoreactivity and hallmark of pRD, we measured the level of neutralizing anti-IFNα autoantibodies (*4*, *34*). All except two patients had high neutralizing antibody titers and concomitant history of CMV exposures **(Fig5R)**. Among all RAG-extrinsic and intrinsic factors, CMV exposure had the highest correlation with the presence of neutralizing anti-IFNα autoantibodies **(Fig5S)**.

### T immune subsets composition, total TCR repertoire and clinical severity in pRD

Next, we dissected the T cell immunophenotype **(FigS3A)** and TCR repertoire in pRD. Our cohort displayed T cell lymphopenia, including total count, CD4+, CD8+ subsets, and naive and recent thymic emigrants across all ages typical of pRD **(Fig6A-B, FigS3B-C)**. As a shared RAG-intrinsic feature, all pRD patients had near absence of TCRVα7.2^+^ mucosal-associated invariant T cells (MAIT) **(Fig6C)** and TCRVα24^+^ invariant natural killer T cells (iNKT) **(FigS3D)**, as previously established (*11*). As a novel feature in T cell biology, Tfh cells expanded and correlated with IDDA2.2 scores in pRD **(Fig5P-Q)**.

**Figure 6.**
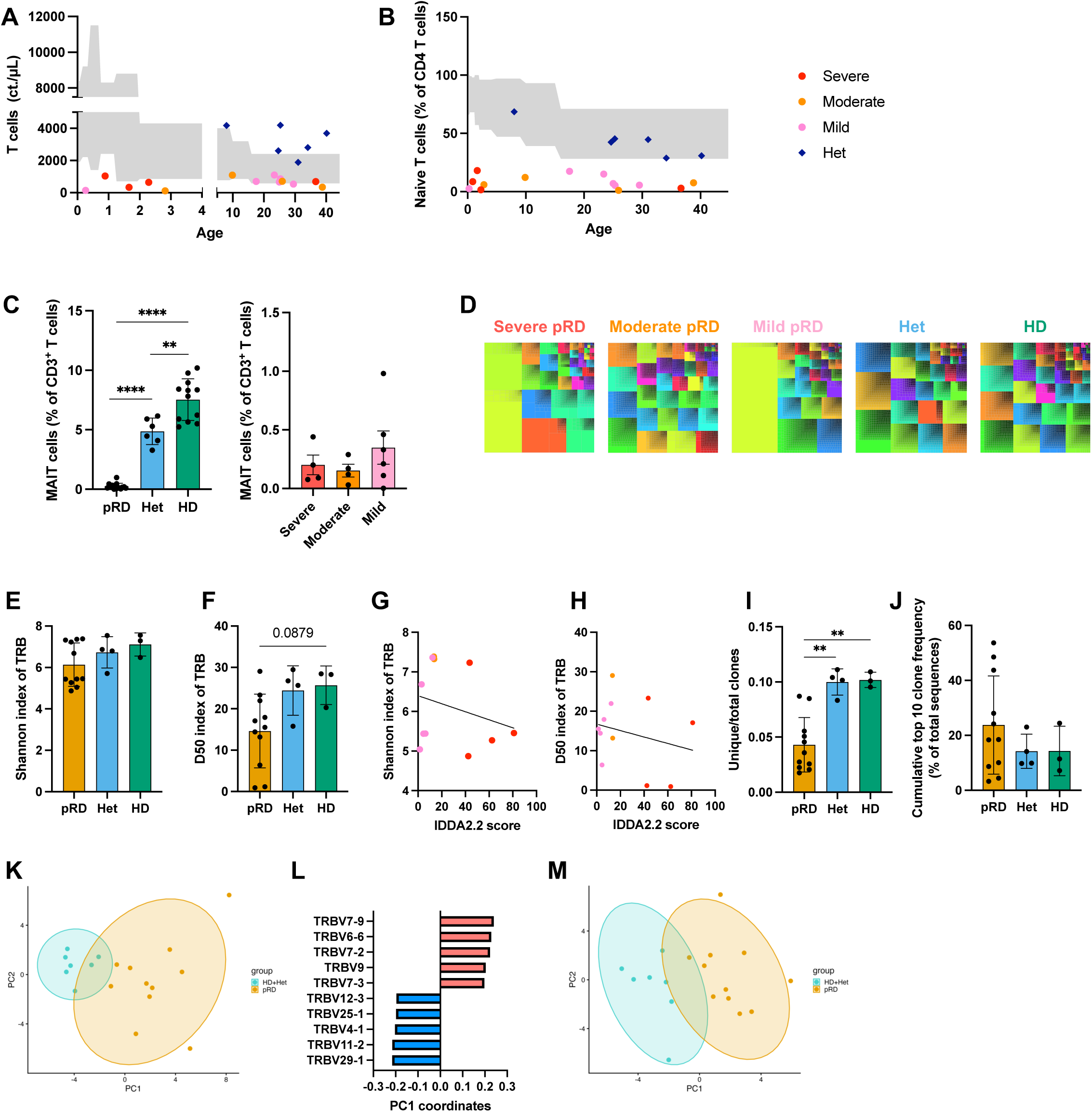
Total CD3+ TCR repertoire indices does not correlate with disease severity in pRD. Immunophenotyping of 14 timepoints from 12 pRD patients and 6 Hets, **A)** CD3^+^ T cell count **B)** CD4^+^ naïve T cell count in pRD by severity clusters, Het and normal range **C)** Frequency of TCRVα7.2+ MAIT cells in pRD, Het and HD **(left panel)** and in pRD patients by severity clusters **(right panel)**. **D)** Treemaps of TRBV gene frequency (top 2500 clones) weighted by the clone size. **E)** Shannon index. **F)** D50 index. **G)** Correlation between Shannon index and IDDA2.2 scores. **H)** Correlation between D50 index and IDDA2.2 scores. **I)** Frequency of unique clones. **J)** Cumulative top 10 clone frequency. **K)** PCA plot for unique TRBV gene segments. **L)** Top 10 unique TRBV gene segments contributing to the separation along PC1. **M)** PCA plot for weighted TRBV gene segments to clone size. One patient was excluded because the raw input sequences were too low (n=50). P≥ 0.05 was considered statistically significant by Mann-Whitney test. Correlation and p value was calculated by Spearman *ρ* and *p* test. Abbreviations: MAIT: Mucosal Associated Invariant T cells

Analysis of the TCR repertoire revealed a trend for lower diversity in pRD as visualized by tree maps **(Fig6D)** and Shannon **(Fig6E)** and D50 **(Fig6F)** indices compared to Hets and HDs. Correlation between diversity indices and IDDA scores showed decreasing tendency **(Fig6G-H)**. Unlike in the case of the BCR, TCR diversity did not change with age **(FigS3E-F)**. In parallel with reduced D50 index, the frequency of unique clones demonstrated a marked decrease in pRD patients **(Fig6I)**, indicating clonotypic expansions, however this did not correlate with the disease severity **(FigS3G)**. Confirming the clonotypic expansion, the frequency of the top 10 clones was elevated in pRD patients **(Fig6J)** although not significantly. This also did not correlate with disease severity **(FigS3H)**.

Analysis of the frequency of individual *V* gene segments **(FigS3I)** showed clear separation along PC1 between the merged group of HDs and Hets and pRD patients **(Fig6K)** consistent with a previous work (*28*). Top 10 PC1 loadings showed differential usage of several *TRBV* segments **(Fig6L)** however disease severity did not separate with the gene usages in pRD patients **(FigS3J)**. Since pRD patients are more susceptible to infections due to decreased TCR/BCR repertoire diversity, we checked for skewing in the *TRBV* gene segments’ usages weighted by the clone size in pRD patients. We found a clear separation between pRD patients and merged HD and Het groups in the utilization of *TRBV* genes **(Fig6M)**, however there was no separation by the IDDA clusters reflecting disease severity **(FigS3K)**.

With these findings we confirmed that unlike B cells, clonal restriction of total TCR repertoire is dominantly defined by partial recombinase activity and none of the indices correlate with clinical severity.

### Variability of T cell competence upon CMV exposure in pRD

Given a shared restricted TCR repertoire within our cohort but divergent clinical outcomes and an IFNγ-driven chemokine signature relating to clinical state, we searched for external factors which could contribute to severe clinical state and trigger T cell dysfunction. Patients with variants of RAG deficiency are highly susceptible to Herpesviruses, including CMV in form of chronic viremia or severe viral disease (*13*, *14*). In physiological settings, adaptive immunity, including the CMV-specific T cells and production of neutralizing antibodies against multiple CMV proteins fully control CMV and sustain latency (*35*). In our pRD cohort and previous studies, we gathered evidence that CMV exposure had the highest correlation with the presence of neutralizing anti-IFNα autoantibodies **(Fig5S)** (*36*). Therefore, we considered CMV exposure as a candidate extrinsic trigger to worsen clinical disease severity in pRD.

Nine of the 15 pRD patients were confirmed of CMV-exposure by detection of viremia and/or the presence of CMV-specific IgG and /or IgM in IgRT naïve cases. We also had access to patient samples before and during severe acute CMV disease (P1, P2). Besides CD8^+^ T cells, γδT cells are also known to expand in acute Herpesviridae infection (*13*). Accordingly, peripheral blood immunophenotyping showed expanded CD8^+^ **(Fig7A)** and γδ T cells **(Fig7B)** in CMV-exposed vs CMV-non-exposed pRD patients and emerged in the time course of P1 and P2 pre and during acute CMV disease **(Fig7A-B)**. CD8^+^ and γδ T cell expansions were more robust in CMV-exposed pRD patients vs Hets and HDs, whereas frequency of CD4^+^ T cells decreased in CMV-exposed patients **(FigS4A)**.

**Figure 7.**
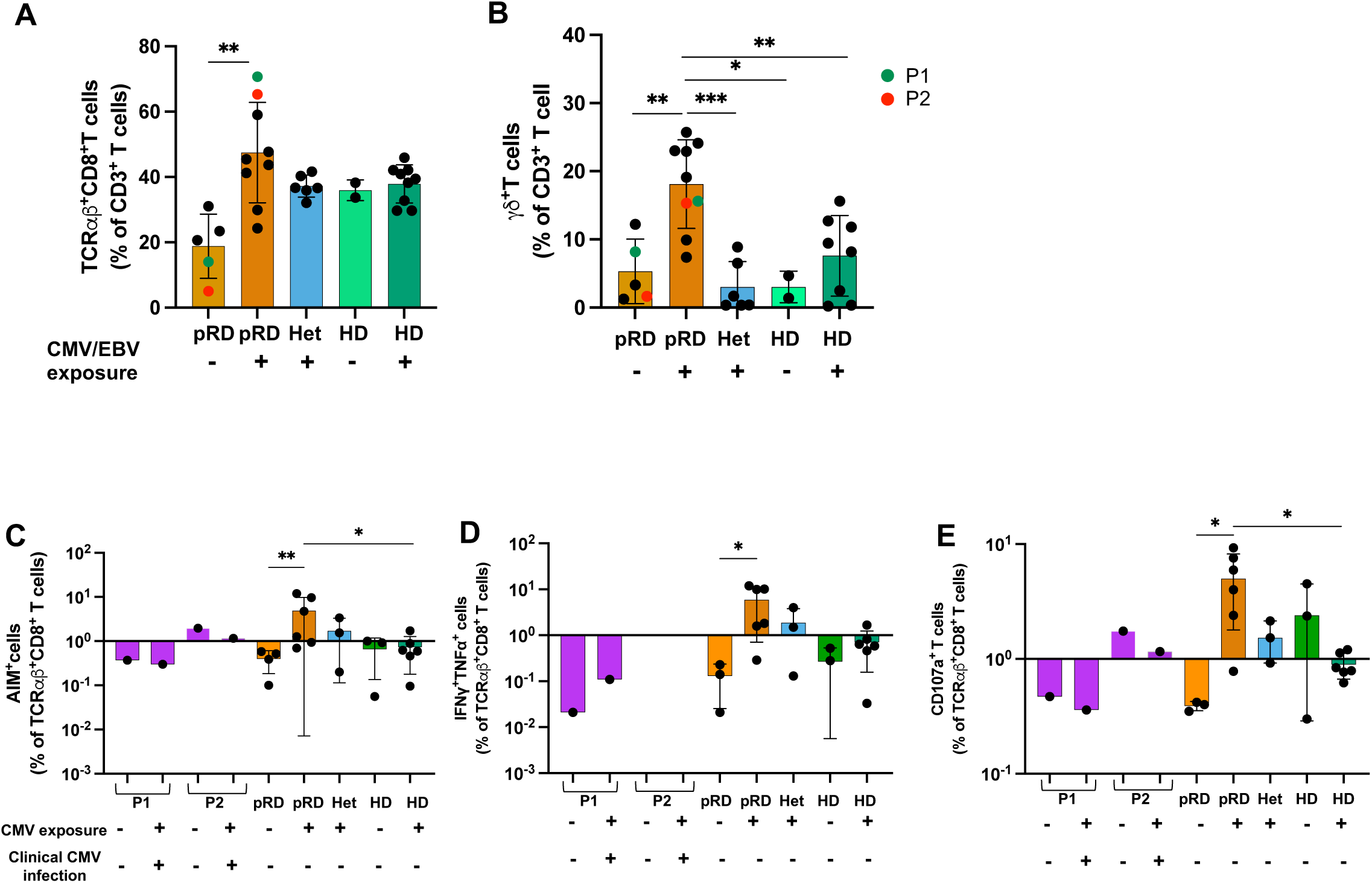
Acute CMV disease ameliorates functional CD8^+^ T cell responses. **(A)** TCRαβ^+^CD8^+^ frequency and TCRγδ^+^ T cell ratio **(B)** in whole blood of pRD timepoints (n=14), Hets (n=6) and HDs (n=12) segregated by Herpesvirus exposure. P1 and P2 timepoints comparing pre- and post-CMV disease are color-coded. **C)** AIM assay comparing CMV stimulated T cells. **D)** Cytokine secretion by CMV peptide stimulated CD8^+^AIM^+^ T cells. **E)** Degranulation capacity of CD8^+^AIM^+^ T cells upon CMV peptide activation. Significance was calculated using Mann-Whitney tests; p≥ 0.05 was considered statistically significant. Abbreviations: AIM: Activation-Induced Marker; CMV: cytomegalovirus; EBV: Epstein-Barr Virus.

To have a detailed look at how history of CMV exposure vs acute disease affects antigen-specific T cell responses in pRD, we set up an *in vitro* assay, where PBMCs were exposed to (1) pan-T cell stimulation (anti-CD3/CD28) to investigate global T cell responses or (2) CMV peptide pool to study CMV-specific T cell responses. We performed three types of flow-based assays: detection of activation induced marker (AIM) expression, cytokine secretion, proliferation and degranulation **(Supplementary Materials 6)**.

Global T cell function after pan-T cell stimulation was variably preserved in both CD4^+^ and CD8^+^ cells in pRD patients **(FigS4B-C)**. We selected patient samples and controls with history of CMV exposure to investigate CMV-specific T cell responses. After CMV peptide stimulation, AIM marker expression **(Fig7C)**, total IFNγ and TNFα secretion **(Fig7D)** and total degranulation capacity by CD8^+^ T cells **(Fig7E)** were all significantly elevated in CMV-exposed pRD patients as compared to CMV negative pRD patients and also higher than CMV-exposed Hets and HDs. P1 and P2 could not respond with AIM expression **(Fig7C)** and cytokine secretion **(Fig7D)** neither pre-nor during acute CMV infection, whereas CD8 degranulation was partly preserved in P2 vs P1 during acute infection **(Fig7E)**. Overall, our pRD patients could mount specific immune response to CMV, however those with acute CMV disease responded less effectively.

## DISCUSSION

For the first time, we established *RAG1* p.C176F as a missense pathogenic founder variant within the genetically insulated Mennonite community. In addition, this study provides the first functional assessment of the recombination activity of the p.C176F variant, demonstrating its deleterious impact on RAG1 function. This was supported by population genetics, multi-lineage pedigree analysis, *in vitro* functional recombination assay, *in silico* analysis, structural studies and clinical characterization of patients with homozygosity. The enrichment of this variant in the Mennonite population is likely due to a bottleneck event that occurred during their historic transatlantic migration from Europe to North America between 1700 and 1800, along with genetic drift and the tendency to marry within their own religious denomination. As individuals carrying this variant are now widespread across Mennonite communities in several geographic areas of the United States and Canada, sequencing studies should be considered when evaluating Mennonite patients with compatible clinical findings or relevant family histories. Increased awareness, non-invasive Prenatal Testing of carrier parents or genetic testing at birth can complement the SCID-NBS process to identify cases in a timely manner.

Another founder RAG1 variant (p.K992E) has been previously described among Old-Order Amish resulting in T^-^/B^-^ SCID / T^+^B^-^ OS immunological phenotypes (*22*, *37*). Unlike these RAG1 SCID/OS patients, our cohort has B^+^ CID phenotype with the ability to produce immunoglobulins, CMV-specific antibodies and anti-CMV T cell response in a subset of patients. As we have shown, the diagnostic process has improved with the introduction of SCID-NBS screening, however the clinical and immunological decline is variable for as long as decades, which create a dilemma for the initiation of anti-microbial prophylaxis (especially herpes viruses), IgRT and raises the question about who, when and how to transplant.

Previously, only a few case reports have described the variable clinical presentation of the patients with the same variants, even within the same family (*7*, *9*). Our unique cohort of 18 pediatric and adult patients with the same *RAG1* p.C176F variant is the largest kindred to date and showcase a clinical landscape where varying composite of infectious and non-infectious complications accumulated at varying rate. We described the compound effect of RAG intrinsic and extrinsic triggers on progressive breaks in peripheral tolerance from the B cell perspective among patients with partially abrogated recombinase activity and B^+^ immune phenotypes (*4*). Many of these patients survived even to adulthood with late onset disease and need for HSCT (*3*, *20*, *21*).

The *RAG1/2* genes are implicated in maintaining both immune competence (host defense supported by TCR/BCR and consequent antibody diversity) and tolerance (pruning of autoreactive T and B cells centrally and indirectly peripherally (*3*)). Therefore, in case of abnormal RAG activity, the residual immune system will face challenges to counter microbes (immune competence) and filter our autoreactive T and B cells (immune tolerance), which together contribute to progression to infections and/or immune dysregulation.

A multi-omics approach to the real-world experience of the CID phenotype of pRD has been described (*11*). Our unique cohort creates a distilled version of this phenomenon where clinical variability arises from the complex interplay of several critical factors, particularly how intrinsic RAG hypomorphism interacts with extrinsic environmental pressures to shape clinical trajectory.

Regarding RAG intrinsic genotype-imposed constraints, we detected dramatically reduced naïve T cell count (and TRECs) and deceased use of distal elements in the T and B cell receptor repertoires, as reflected by low TCRVα7.2 MAIT cells and low representation of IGHVJ5 and J6, respectively.

Extrinsic drivers of clinical and immunological phenotype likely include the T_H1/_IFNγ-driven inflammatory milieu (CXCL9/10 levels) and acute or chronic exposure to microbial antigens, such as CMV, secondary to inefficient clearance. We also saw predisposition for worse clinical outcome among females, which may be partially explained by augmented innate signals, such as TLR7, that is coded by the X chromosome. CD21^lo^ B cells (co-expressing CD11c and Tbet) in particular are very sensitive to TLR7 stimulation and therefore female gender may predispose pRD patients to B cell immune dysregulation in the extrafollicular space. This phenomenon also included longitudinal decline of transitional B cells, expansion of activated naïve B cells, and elevated anti-IFNα autoantibodies (*4*, *15*, *29*).

This joint contribution of intrinsic and extrinsic drivers creates a progressive decline in the ability of T cells to eliminate novel or chronic existing triggers (e.g. CMV viremia) and divert proper B cell responses with BCR diversification and antibody maturation from the germinal centers to the extrafollicular space. The crosstalk of expanded Tfh cells and ABCs in the T_H1 i_nflammatory milieu of the extrafollicular space may promote selection of autoreactive prone early B cells (VH4.34 BCR) and their fast transition to antibody secreting plasma cells, giving rise to autoantibodies (e.g. anti-IFNα, 9G4) (*29*). We reconfirmed correlation of history of CMV exposure with presence of neutralizing anti-IFNα autoantibodies (*36*).

The multiomics assessment from our cohort demonstrates that both sexes are vulnerable for severe viral infections such as CMV in early childhood likely related to restricted TCR repertoire and in the setting of grossly preserved B cell compartments and function. Regarding antigen-specific responses, T cells from pRD patients are capable of responding to CMV, albeit to at various extents: a subgroup of patients lacks or partially lacks T cell competence and consequently experiences severe CMV clinical disease as opposed to those who retained competence with no progression of clinical disease. With environmental triggers (e.g. viral infections), B cell counts and function decline in pRD with age.

Regarding implications for risk for worsening clinical course, RAG-intrinsic factors poorly discriminated which patient in our cohort has clinical decline. In contrast, severity scores correlated with many of the factors driven by extrinsic processes: we noted correlation with severity of clinical disease and serum factors reflecting T_H1/_ IFNγ-driven pathology (CXCL9/10), cellular (Tfh expansion) and B cell receptor repertoire level (relative contribution of BCR VH4.34).

Longitudinal sampling of a cohort of patients sharing the same *RAG1* variant and assessment of their clinical severity at multiple timepoint provided a new dimension of timing for infectious predisposition and dysregulation in pRD. Our findings have clear implications that could improve decision making and patient care. We have provided multiple layers of data for key conceptual insights into disease trajectory, level of immune dysregulation, the presence of viral modulation and therapeutic timing. These biomarkers along with IDDA scoring and conventional immune evaluation could contribute to the complicated decision-making framework used by physicians (Immunologist, Transplant Hematologist/Oncologist) and families to consider prognosis, monitoring and intervention, including anti-viral prophylaxis, IgRT or HSCT.

Our study has several limitations. First, the current version of IDDA score does not account for the relative contribution of different clinical manifestations. Further optimization of this score, especially for pRD patients, is required. Second, there are no universal prognostic biomarkers for pRD and the clinical outcome of an individual is influenced by a combination of environmental triggers and therapies. The role of background HLA haplotype, antigen load, TCR repertoire, and inflammatory responses need to be assessed prospectively, which will be addressed in future studies.

Another limitation of our study is the relatively modest absolute cohort size, which is expected for a rare monogenic disorder. Although our findings do not provide an in-depth molecular characterization of disease pathogenesis, a major strength of this study is the establishment of a comprehensive analytical framework that integrates a broad range of cellular and immunological parameters with clinical phenotypes, thereby facilitating translation to clinical practice and providing mechanistic human relevance to the recent findings by Hong et al. (*25*). Importantly, this represents the largest cohort reported to date of patients carrying the identical RAG1 variant. However, despite this genetic homogeneity, the cohort exhibits substantial phenotypic heterogeneity, and the sample size remains insufficient to broadly generalize these findings. Nevertheless, parallel studies in other monogenic immunodeficiencies, including NFKB1, CTLA4, and APDS (*38*, *39*), have reported similar patterns of early B-cell activation and dysregulated extrafollicular Tfh–B-cell crosstalk, supporting the broader relevance of these observations (*38*, *39*).

In conclusion, pRD presents with a diverse clinical and immunologic profile. This is a result of a complex interplay of intrinsic RAG hypomorphism with extrinsic environmental pressures to shape clinical trajectory. These pRD patients require regular clinical evaluation to monitor the development of immune dysregulation and assess disease-related immunologic biomarkers. Prospective studies are needed to further identify who should undergo HSCT and when it should be performed as the definitive therapy.

## Materials and Methods

### Study Design

This is a multi-center observational translational cohort study to characterize the clinical and immunologic heterogeneity of patients with homozygous RAG1 c.527G>T (p.Cys176Phe) variant of uncertain significance within a genetically and environmentally homogeneous Old Order Mennonite population. The primary objective was to confirm the pathogenicity of the RAG1 variant, determine whether individuals sharing identical genotype and ancestry demonstrate divergent clinical trajectories, to identify RAG-intrinsic and extrinsic triggers and candidate immunologic biomarkers of disease severity. Pre-specified hypotheses were that (i) RAG1 c.527G>T (p.Cys176Phe) is a pathogenic variant, (ii) substantial clinical variability as measured by the Immune Deficiency and Dysregulation Activity score (IDDA2.2) would be observed despite shared genotype (*26*), (iii) ability to distinguish shared RAG-intrinsic features of immune abnormalities (iv) identify extrinsic triggers and related immune abnormalities and (v) viral-specific immune competence, particularly against herpesviruses, would associate with clinical severity and outcomes.

Eighteen individuals homozygous for RAG1 c.527G>T were enrolled, along with 11 family controls (heterozygous carriers and wild-type relatives). For final clinical outcome analyses, 15 patients were included. Three patients were excluded prospectively from clinical severity analyses because an independent second monogenic disorder substantially altered their phenotype and medical management. Participants were recruited under protocols approved by the Institutional Review Boards of the University of South Florida (Pro00035468; Principal Investigator: J.E.W.) and the Clinic for Special Children (CFSC 2008-095; Principal Investigator: K.W.B.). The study was conducted in accordance with institutional guidelines and the Declaration of Helsinki. Written informed consent was obtained from all adult participants or from the legal guardians of minors. Inclusion criteria were defined prior to analysis and required Mennonite ancestry and molecular confirmation of homozygous RAG1 c.527G>T. Exclusion criteria included the presence of an additional independent genetic diagnosis or insufficient clinical documentation.

Sample size was determined by the number of patients, heterozygotes, and healthy donors (HDs) included in the cohort, as well as the number of available clinical assessment time points per patient. Given the retrospective nature of the study, follow-up duration was not predefined; instead, we aimed to include the most recent and complete data for all individuals. As dates of birth, diagnosis, and hematopoietic stem cell transplantation (HSCT) varied across patients, the length of follow-up differed between individuals.

This was a non-randomized observational study. Participants were enrolled consecutively based on genotype confirmation. Laboratory personnel performing functional and biomarker assays were blinded to clinical severity categories when feasible. Clinical scoring using IDDA was performed independently of laboratory measurements.

It should be noted that some patients changed IDDA cluster classification during their disease course and could belong to different clusters at different clinical assessment time points; for example, patient P14 was classified as mild after pregnancy. Accordingly, depending on whether analyses considered the full disease history or specific time points, the number of cases within each cluster varied.

Clinical data were collected through structured in-person interviews, telephone interviews, comprehensive review of medical records, and direct communication with treating physicians. Demographic characteristics, family history, immunologic parameters, infectious and autoimmune manifestations, and treatment histories were systematically documented. Longitudinal disease burden was quantified using the IDDA scores, applied at initial presentation and across follow-up timepoints.

All relevant data were included in the analyses. One outlier was excluded in **Fig4**. In **Fig3C**, two patients (P1 and P2) were additionally excluded in a secondary analysis due to clinical considerations, as early CMV infection in these cases was deemed likely to confound the results. Analyses were first performed including all patients and subsequently repeated excluding P1 and P2, as this approach provided a more clinically meaningful interpretation.

Primary and secondary endpoints were defined based on key milestones of disease course, including dates of birth, diagnosis, HSCT, and death. The number and type of experimental replicates are detailed in the Materials and Methods section according to the specific assays used.

### Experimental design

Laboratory experiments were performed using biological replicates per participant and internal technical controls in accordance with manufacturer and previously validated protocols. Where feasible, assays were conducted in duplicate or repeated under standardized conditions to confirm reproducibility. Outliers were not excluded unless attributable to documented technical failure, including assay control failure or acquisition error. No biological data points were excluded solely on the basis of statistical extremity. Number of samples in the experiments was dependent on sample availability at that time. Sample and replicate numbers are indicated both in detailed methods description and in figure legends.

### Statistical analysis

Individual-level data are presented in data file S1. Number of samples were determined by the number of timepoints of clinical assessment, and sample availability. The statistical tests used are specified in the figure legends. Pairwise comparisons were performed by Mann-Whitney U test (one variable). Categorical variables were analyzed by Fisher’s exact test. Correlations were assessed by the non-parametric Spearman rho test. *p* < 0.05 was considered statistically significant. K-means clustering of IDDA scores was performed in R, and the results were visualized using the *ggplot2* package (*40*). Statistical analyses were performed using GraphPad Prism v.10.4.1. All data are presented as means ± SD.

## Supporting information

Combined Supplementary Materials

## Data Availability

All data produced in the present study are available upon reasonable request to the authors

## Acknowledgements

Research laboratory studies were performed samples under IRB approved protocols at University of South Florida (USF-Pro00035468 (PI, JEW) and NIH IRB-approved protocol 18-I-0041 (NCT03394053). We also gratefully acknowledge the SCID Angels For Life Foundation and the late Heather Smith for her dedication and invaluable contributions.

## Author contributions

E.P., M.To., E.S.J.E., M.N.T., L.D.N., K.A.S., and J.E.W. designed the studies. E.P., M.To., P.R.S., M.Ta., K.A.L., D.J.G., P.B., M.Bo., A.C.N., E.B.O.J., R.S., T.J.T., B.U., K.C., Y.N.L., M.Bu., A.E.K., E.G.P., L.D.N., and J.E.W. performed the experiments. E.P., M.To., P.R.S., M.Ta., Z.G., K.A.L., D.J.G., P.B., M.Bo., A.C.N., G.H., A.H., E.B.O.J., M.S.O., O.O., M.S., C.D.S., S.O.S., R.S., T.J.T., B.U., K.C., Y.N.L., M.Bu., A.E.K., E.Pu., L.D.N., and J.E.W. analyzed the results. E.P., P.R.S., R.M., P.M., J.J.B., S.C., D.D., J.H., S.E.H., K.E.H., A.J., M.K., K.P., J.P.R., C.M.S., J.W.V., B.R.W., J.W., W.Z., M.Y., N.L.R., L.P., K.W.B., L.D.N., K.S., and J.E.W. participated in acquisition of data. E.P., M.To., P.R.S., M.Ta., Z.G., P.B., Ma.B., I.K.C., A.C.-N., O.D., E.S.J.E., G.H., A.H., E.C.M., E.B.O.J., M.S.O., O.E.O., M.S., C.D.S., S.O.S., R.S., M.Te., Y.Z., S.X., M.Bu., A.E.K., N.L.R., E.G.P., L.D.N., K.A.S., and J.E.W. interpreted the results. E.P., M.To., P.R.S., Z.G., S.X., A.E.K., E.G.P., and J.E.W. wrote and edited the manuscript. E.P. and M.To. contributed equally as shared first authors. L.D.N., K.A.S., and J.E.W. contributed equally as shared senior authors.

## Data, code, and materials availability

All data associated with this study are in the paper or Supplementary Materials.

