## Supplementary material for "Multi-omics approach to map a clinically diverse large cohort with homozygous founder RAG1 p.C176F variant among Old Order Mennonites": Combined Supplementary Materials

**Supplementary Materials and Methods**

*1. Determination of RAG1 recombination activity*

The recombination activity of human RAG1 p.C176F mutant was assessed using a previously validated flow cytometry-based reporter assay in a *Rag1*$^{-/-}$ pro-B cell lines reconstituted with wild-type or mutant RAG1 in triplicates (*1*). Residual V(D)J recombination activity was quantified relative to wild-type control and expressed as a percentage of wild-type activity.

*2. Whole blood immunophenotyping and PBMC isolation*

Peripheral blood samples were collected and processed within 48 hours into plasma, peripheral blood mononuclear cells (PBMCs), or whole blood. Samples were de-identified, coded, and stored in institutional biorepositories under standardized biosafety protocols.

100 µl from whole blood specimen was washed with staining buffer (2% BSA, 2mM EDTA in PBS) to remove circulating antibodies and stained with fluorochrome-conjugated antibody panels targeting myeloid and B cell subsets (**Table S2, Panel A**). 100 µl from whole blood specimen was stained directly with the antibody cocktail for T-cell immunophenotyping (**Table S2, Panel B**). After multiple washing steps with staining buffer and red blood cell lysis (10x FACS lysing solution, BD Biosciences), counting beads (Spectrosort from Cytek or 123 beads from ThermoFisher or Precision Count beads from BioLegend) were added to the samples. Stained cells were acquired with either a BD LSR II platform or Cytek Northern Lights spectral flow platform. Immunophenotyping data were visualized with FlowJo software v.10.8.1 (TreeStar).

Rest of the blood was used for isolating PBMCs using Ficoll-Paque PLUS (Cytiva). Cells were stored at -80°C and liquid $N_2$ for further use.

*3. Measurement of soluble plasma inflammatory biomarkers and lipid mediators*

A total of 20 soluble inflammatory biomarkers were measured in plasma samples from patients with pRD (n=13), heterozygous carriers (n=5), and healthy controls (n=11), using Legendplex™ cytometric bead array (BioLegend) following the manufacturer instructions in duplicates, incorporating both the 13-plex Human Essential Immune Response Panel (IL-4, IL-2, CXCL10/IP10, IL-1β, TNFα, CCL2/MCP-1, IL-17A, IL-6, IL-10, IFNγ, IL-12p70, CXCL8/IL-8, TGFβ1) and a customized 7-plex panel (CXCL9, IFNα, IL-18, IL-13, BAFF, CCL17, CCL22). P1 T2 sample's one replicate was excluded from CCL17 and CCL22 data due to saturation; P8 was excluded from BAFF data due to rituximab treatment. Data analysis was performed using Data Analysis Software Suite for Legendplex v1. Graphic visualization was conducted with GraphPad Prism v.10.4.1 (GraphPad Software, Boston, MA) and R package *fmsb* for spider plotting. CXCL9, CXCL10 and IFNγ concentrations were confirmed with ELISA (R&D Systems) following the manufacturer's recommendations.

For detecting lipid mediators from the plasma samples of ten pRD patients (severe=4, moderate=2, mild=4), five RAG1 heterozygotes and five HDs, we used high performance liquid chromatography/mass spectrometry (HPLC/MS) system. For preparing the plasma samples, methanol was used for protein precipitation. After a centrifugation step (15,000g, 10 min, 4°C), supernatant was removed and placed in a Stuart block heater for evaporation. The sample were reconstituted with 1:1 water : methanol mix and the supernatant was centrifuged (15,000g, 10 min, 4°C), was transferred to HPLC vials and injected to the LC-MS (Sciex 6500+ Qtrap). The data integration was performed with Sciex OS2.0. The calibration curve and the raw data was processed as previously described (*2*). Data was analyzed using GraphPad Prism v.10.4.1; heatmap and PCA were plotted using *ggplot2* package in R (*3*).

*4. Measurement of CMV/EBV viral load, viral specific IgG/M, and anti-IFNα autoantibodies*

Plasma CMV and EBV DNA viral loads were measured by real-time quantitative PCR by Clinical Laboratory Improvement Amendments (CLIA)-certified laboratory (Eurofins Viracor Clinical Diagnostics, KS, USA). CMV- and EBV-specific IgG antibodies were detected using enzyme-linked immunosorbent assay (ELISA) (13 pRD patients, 6 Hets and 13 HDs) in our laboratory (MyBiosource). IFNα was screened by in-house developed ELISA in 384-well high-binding MaxiSorp polystyrene microplates with 50 µL per well of Recombinant Human IFN-α2 (carrier-free) (0.5µg/mL in PBS, pH 7.0; BioLegend). Plates were washed three times with PBST containing 0.05% Tween 20 (PBST) and blocked for 2 hours at room temperature with 80 µL per well of 5% BSA-PBS. After washing, 50 µL of patient or healthy control plasma samples (serially diluted starting at 1:100 to 1: 1638400 in 1% BSA-PBS) were added in duplicate and incubated for 1.5 hours at room temperature. Plates were washed three time with PBST, followed by a 45-minute incubation with 50 µL per well of horse radish peroxidase conjugated anti-human IgG (diluted 1:5000 in 1% BSA-PBS; Invitrogen ThermoFisher Scientific). After three additional washes, 50 µL per well of 3,3",5,5'-tetramethylbenzidine substrate was added for 5 minutes in dark, and the reaction was stopped with 50 µL of 1N $H_2SO_4$. Absorbance was measured at 450 nm with 570 nm background correction using a VANTAstar microplate reader (BMG Labtech). The titers were determined by serial dilution, as described previously (*4*). The cutoff for positivity was defined as the mean plus three standard deviations (Mean + 3SD) of the values from healthy controls.

*5. Measurement of neutralization capability of anti-IFNα autoantibodies*

Patients who tested positive for anti-IFNα autoantibodies were further analyzed via a flow-based neutralization assay as detailed *(5)*. Briefly, healthy donor frozen PBMCs were thawed, counted and aliquoted at 350,000 cells/well. Separately, recombinant IFNα was exposed to patient or HD plasma to allow for antibody neutralization of cytokines. PBMCs were mixed with the plasma and IFNα solution for a final concentration of 10ng/mL and 10% plasma and allowed to incubate at 37 °C for 15 minutes to allow for STAT1 phosphorylation. Afterwards, cells were fixed and stained for CD14 and pSTAT1 and run on the flow cytometer.

*6. Virus-induced T cell activation induced marker/intracellular cytokine production assays*

Patient PBMCs were thawed and plated at 1,000,000 cells/mL in U-bottom tissue-culture treated plates (Costar); a small aliquot was stained with live–dead discriminator (1:1000, Fixable Viability Dye eFluor 780, ThermoFisher Scientific). Cells were then allowed to rest overnight at 37°C. After resting, cells were then exposed to one of three conditions: [1] unstimulated (dimethyl-sulfoxide at 0.5%), [2] 7.05 µg/mL anti-CD3 and 6.6 µg/mL anti-CD28 antibodies (1:200 both from BioLegend) [3] 1 µg/mL CMV-peptide mix (JPT Peptide Technologies). 2 µM Monensin and 5 µg/mL Brefeldin A (both from BioLegend) were added at the time of stimulation to prevent cytokine secretion. Cells were incubated for 16 hours at 37°C. Then cells were stained for viability and fixed in 2% paraformaldehyde. Cells were stained in perm/wash buffer (BioLegend) with an antibody cocktail. Detection of activation induced marker (AIM) expression (CD69/CD134 for $CD4^+$ T cells and CD69/CD137 for $CD8^+$ T cells), cytokine secretion (IFNγ, TNFα and IL-2). To detect degranulation of $CD8^+$ T cells, we included CD107a (LAMP1) in our panel (**Table S2, Panel AIM/ICS/CTV**).

To assess proliferation, cells were labeled with CellTrace Violet (ThermoFisher Scientific), then plated and stimulated as above, but without Brefeldin A or Monensin, and allowed to proliferate for 5 days. On Day 5, cells were fixed, stained, and analyzed as above for T cell subsets and exhaustion markers (**Table S2, Panel AIM/ICS/CTV**).

Data was acquired on a Cytek Northern Lights spectral flow platform. Data was analyzed with FlowJo software v.10.8.1 and visualized with GraphPad Prism v.10.4.1.

*7. Bulk BCR and TCR sequencing*

Genomic DNA was extracted from whole blood without lymphocyte enrichment. The *TRB* and *IGH* repertoire libraries were generated from 150 ng of genomic DNA extracted from the patients' and healthy controls' peripheral blood using primers for conserved regions of *V* and *J* genes, according to manufacturer's protocol (LymphoTrack, Invivoscribe Technologies, France and USA). The resulting libraries were purified using AMPure XP beads (Beckman Coulter, USA) and were quantified using Qubit DNA quantification assays (Qubit, Thermo Fisher Scientific, USA). Furthermore, quality control assessment of the libraries was carried out using the TapeStation system (DNA ScreenTape Analysis, Agilent Technologies, USA). Next, the libraries were pooled and sequenced using Mi-Seq Illumina technology (Illumina Inc, USA).

For nucleotide sequence processing and annotation, We used a slightly modified version of the airrmine pipeline (*6*) (https://github.com/blazsop/airrmine), which will be described in the following sections. The resulting .fastq files from sequencing were analyzed with IMGT/HighV-QUEST v1.9.6 (*7*), querying for human B heavy chain (IGH) and T cell receptor beta chain (TRB) sequences. The resulting AIRR-formatted files containing the sequence annotations were parsed using Change-O and its function MakeDb.py v1.3.4 with "extended" annotation columns (*8*). Non-productive sequences were removed early by the airrmine pipeline. Within each sample, we counted identical sequences to reduce redundant records.

To account for PCR errors and mitigate inflation of clone diversity, we used the airrmine pipeline to identify near-identical sequences. The identification was achieved via Hamming distance calculation with the seqDist function (alakazam v1.1 (*8*)). Abundant sequences (four sequences or more) were identified, and any sequence with distance ≤ from an abundant sequence were collapsed to that abundant sequence.

For clonal assignment, we calculated the distribution of pairwise Hamming distances among sequences for each sample to identify the split point between within-clone and between-clone distances. We visually inspected the histograms to modify the airrmine automatically set threshold when needed. The automatically set and final distance thresholds are available as ...... The thresholds were used by airrmine as an argument to the DefineClones.py v1.3.4 function from Change-O, by grouping based on V, J, and CDR3 sequences.

We reconstructed germline sequences per clone using Change-O (CreateGermlines.py v1.3.4 function). We provided IGH human reference germline sequences, which are included in the original airrmine pipeline. For TRB, we obtained reference sequences from IMGT (https://www.imgt.org/vquest/refseqh.html). For B-cell IGH sequences only, somatic hypermutation (SHM) load was quantified on clonal, germline-paired sequences using SHazaM (*8*) as implemented in airrmine. Mutation counting was achieved also with SHazaM v1.0.2 by calculating its BASELINe (*9*) parameter.

Per-sample repertoire diversity was summarized with airrmine from clone AIRR tables. The Shannon-Wiener, Simpson and D50 indexes were calculated. We also generated V-gene summary visualizations (a.k.a. treemaps) using the treemapify v2.5.6 (*10*) and ggplots v3.5.1 (*3*). To reduce treemap complexity, we downsampled the data set to the top 2,500 most abundant clones. Additionally, we generated AIRR networks with the airrmine pipeline and the igraph package v2.2.1.

The nucleotide sequences are available at SRA. The resulting IMGT annotations are available at Zenodo. Data analysis was performed using the airrmine pipeline (https://github.com/blazsop/airrmine) with modifications as outlined in this GitHub repository: https://github.com/oospina/BCR_TCR_Repertoire_Immune_Deficiency.


1. Y. N. Lee, F. Frugoni, K. Dobbs, J. E. Walter, S. Giliani, A. R. Gennery, W. Al-Herz, E. Haddad, F. Ledeist, J. H. Bleesing, L. A. Henderson, S. Y. Pai, R. P. Nelson, D. H. El-Ghoneimy, R. A. El-Feky, S. M. Reda, E. Hossny, P. Soler-Palacin, R. L. Fuleihan, N. C. Patel, M. J. Massaad, R. S. Geha, J. M. Puck, P. Palma, C. Cancrini, K. Chen, M. Vihinen, F. W. Alt, L. D. Notarangelo, A systematic analysis of recombination activity and genotype-phenotype correlation in human recombination-activating gene 1 deficiency. *Journal of Allergy and Clinical Immunology* **133** (2014).

2. G. Upadhyay, L. Duong, V. Kain, M. Ghobrial, M. K. Marimuthu, A. Fouad, T. J. Yeatman, J. D. Herazo-Maya, G. V. Halade, Cannabis cigarette smoking disrupts mice multi-organ bioactive lipid metabolism and inflammation-resolution signaling in an obesogenic setting. *Life Sci.* **383**, 124076 (2025).

3. H. Wickham, “Data Analysis” (2016), pp. 189–201.

4. J. E. Walter, L. B. Rosen, K. Csomos, J. M. Rosenberg, D. Mathew, M. Keszei, B. Ujhazi, K. Chen, Y. N. Lee, I. Tirosh, K. Dobbs, W. Al-Herz, M. J. Cowan, J. Puck, J. J. Bleesing, M. S. Grimley, H. Malech, S. S. De Ravin, A. R. Gennery, R. S. Abraham, A. Y. Joshi, T. G. Boyce, M. J. Butte, K. C. Nadeau, I. Balboni, K. E. Sullivan, J. Akhter, M. Adeli, R. A. El-Feky, D. H. El-Ghoneimy, G. Dbaibo, R. Wakim, C. Azzari, P. Palma, C. Cancrini, K. Capuder, A. Condino-Neto, B. T. Costa-Carvalho, J. B. Oliveira, C. Roifman, D. Buchbinder, A. Kumanovics, J. L. Franco, T. Niehues, C. Schuetz, T. Kuijpers, C. Yee, J. Chou, M. J. Masaad, R. Geha, G. Uzel, R. Gelman, S. M. Holland, M. Recher, P. J. Utz, S. K. Browne, L. D. Notarangelo, Broad-spectrum antibodies against self-antigens and cytokines in RAG deficiency. *Journal of Clinical Investigation* **125**, 4135–4148 (2015).

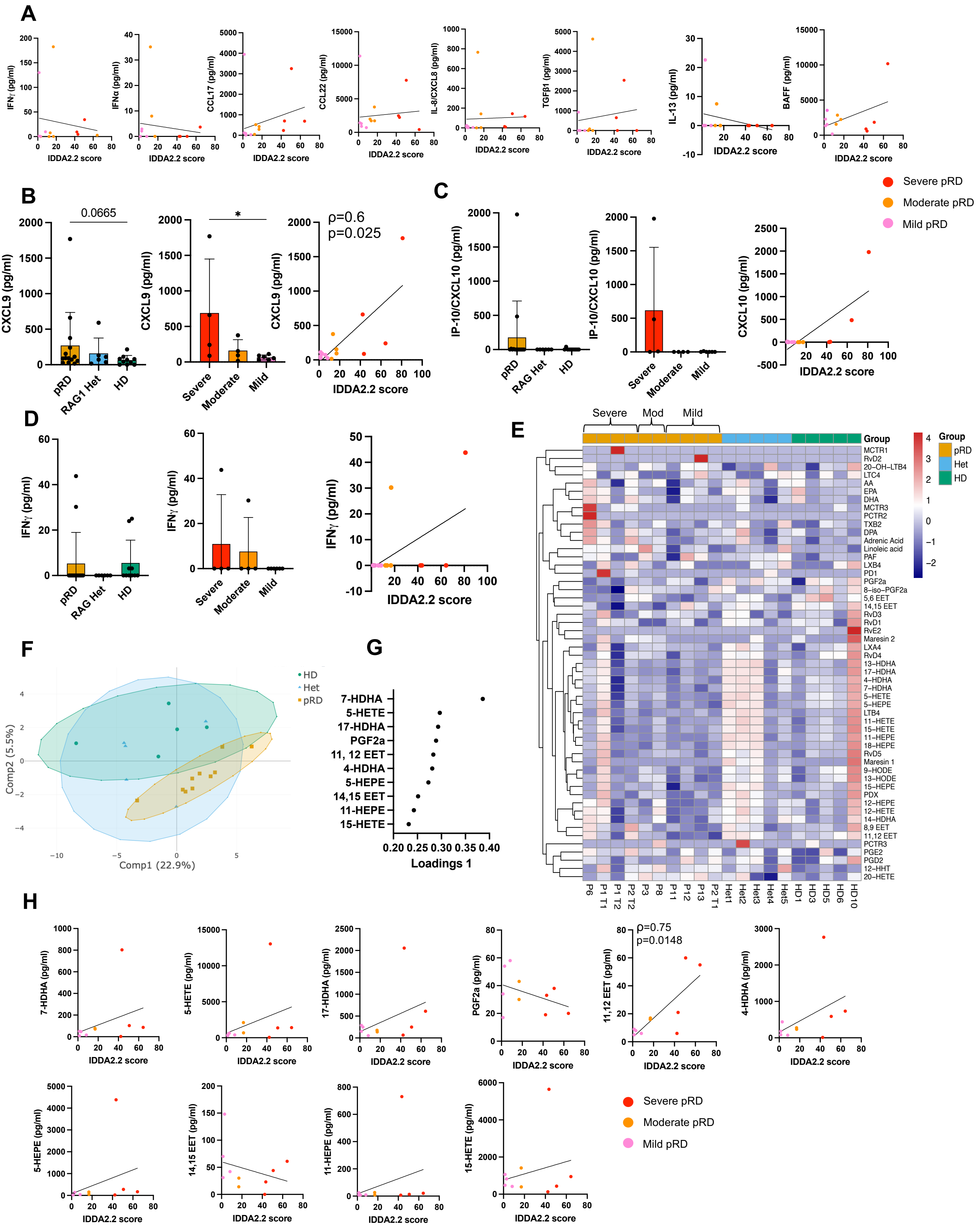


**Figure S1. Cytokines/chemokines highest in the pRD group and lipid mediators are not correlated with disease severity.**
20 plasma factors were measured from sample timepoints from pRD patients (n=13), Hets (n=5) and HDs (n=11). **A)** Eight plasma soluble factors highest in the pRD group from severe (n=4), moderate (n=4) and mild (n=5) patients' timepoints were correlated with IDDA2.2 scores. CXCL9 **(B)**, CXCL10 **(C)** and IFNγ **(D)** concentrations by ELISA from pRD patients (n=14), Hets (n=6) and HDs (n=10) **(left panels)** and from the disease groups (severe=4, moderate=4, mild=6) **(middle panels)**. Their correlation with IDDA2.2 scores were plotted on the **right panels**. **E)** Heatmap showing the concentrations of the individual lipid mediators by pRD patients (n=10), Hets (n=5) and HDs (n=5). Data were $log_{10}$ transformed and scaled. **F)** 2D sparse Partial Least Squares Discriminant Analysis (sPLS-DA) plot showing separation of pRD patients, Hets and HDs. Ellipses represent 95% confidence interval. **G)** Loading plot for Component 1's top 10 lipid mediators. **H)** Correlation between the top 10 mediators for Component 1 and the IDDA2.2 scores.
Significance was calculated by Mann-Whitney test, p≥ 0.05 was considered significant. Correlation and p value was calculated by Spearman correlation and *p* test indicated by the ρ correlation coefficient.

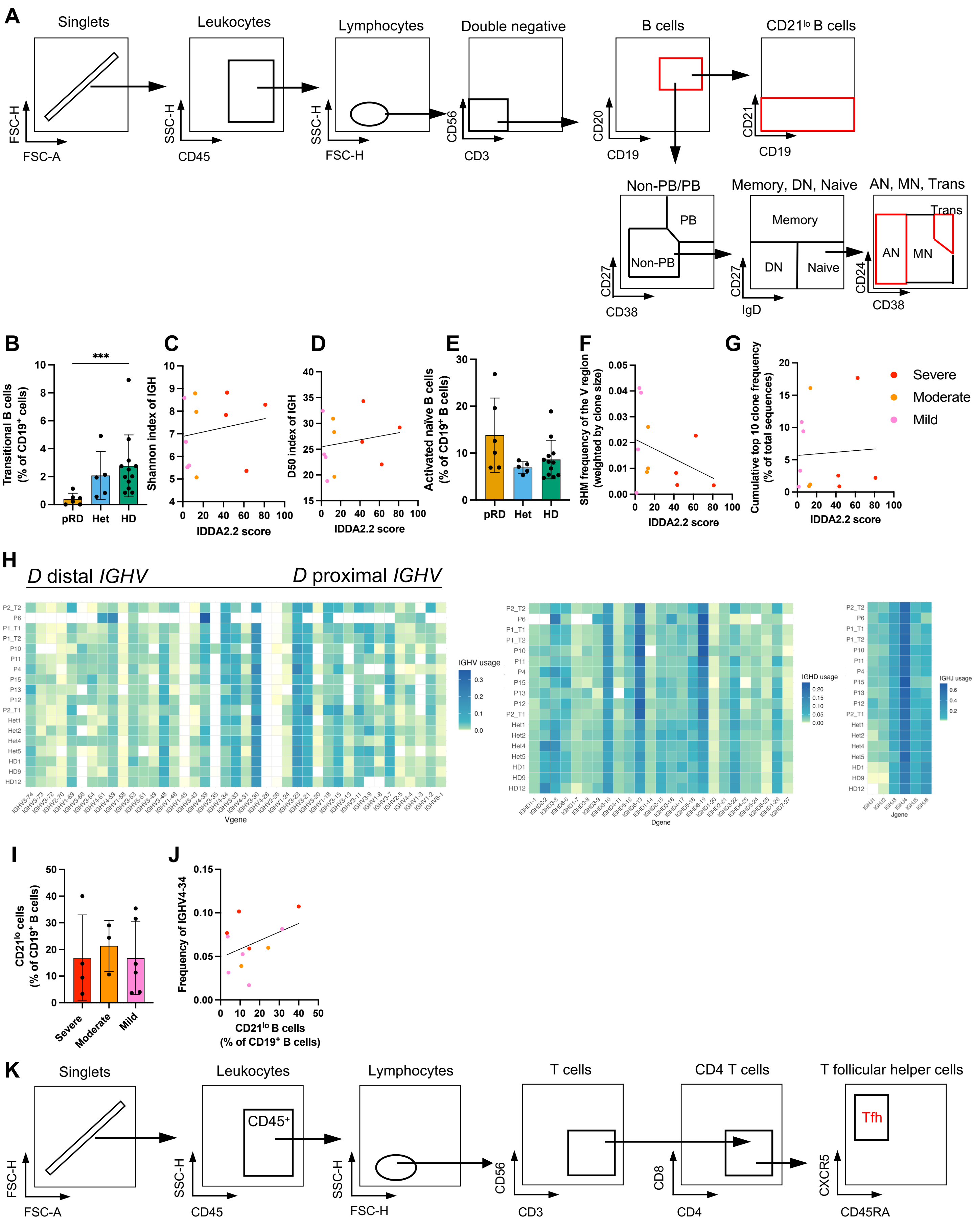


**Figure S2. Whole blood B cell and Tfh immunophenotyping and characteristics of BCR repertoire in pRD. A)** Gating strategy for B cells, transitional, activated naïve (AN) B cells. **B)** Frequency of transitional B cells in pRD patients, Hets and HDs. Correlation between Shannon index **(C)** and D50 index **(D)** and IDDA2.2 scores. **E)** Frequency of AN B cells in pRD patients, Hets and HDs. Correlation between somatic hypermutation (SHM) rate **(F)** and cumulative top 10 clone frequency **(G)** and IDDA2.2. **H)** Heatmaps showing the frequency of IGHV gene segments **(left panel)** from distal to proximal to D region; IGHD gene segments **(middle panel)** and IGHJ gene segments **(right panel)**. **I)** Frequency of CD21$^{lo}$ B cells in pRD patients grouped by disease severity. **J)** Correlation between frequency of IGHV4-34 and CD21$^{lo}$ B cell frequency. **K)** Gating for Tfh cells. Significance was calculated using Mann-Whitney tests; $p \geq 0.05$ was considered statistically significant. Correlation and p value was calculated by Spearman correlation ($\rho$) and $p$ test.

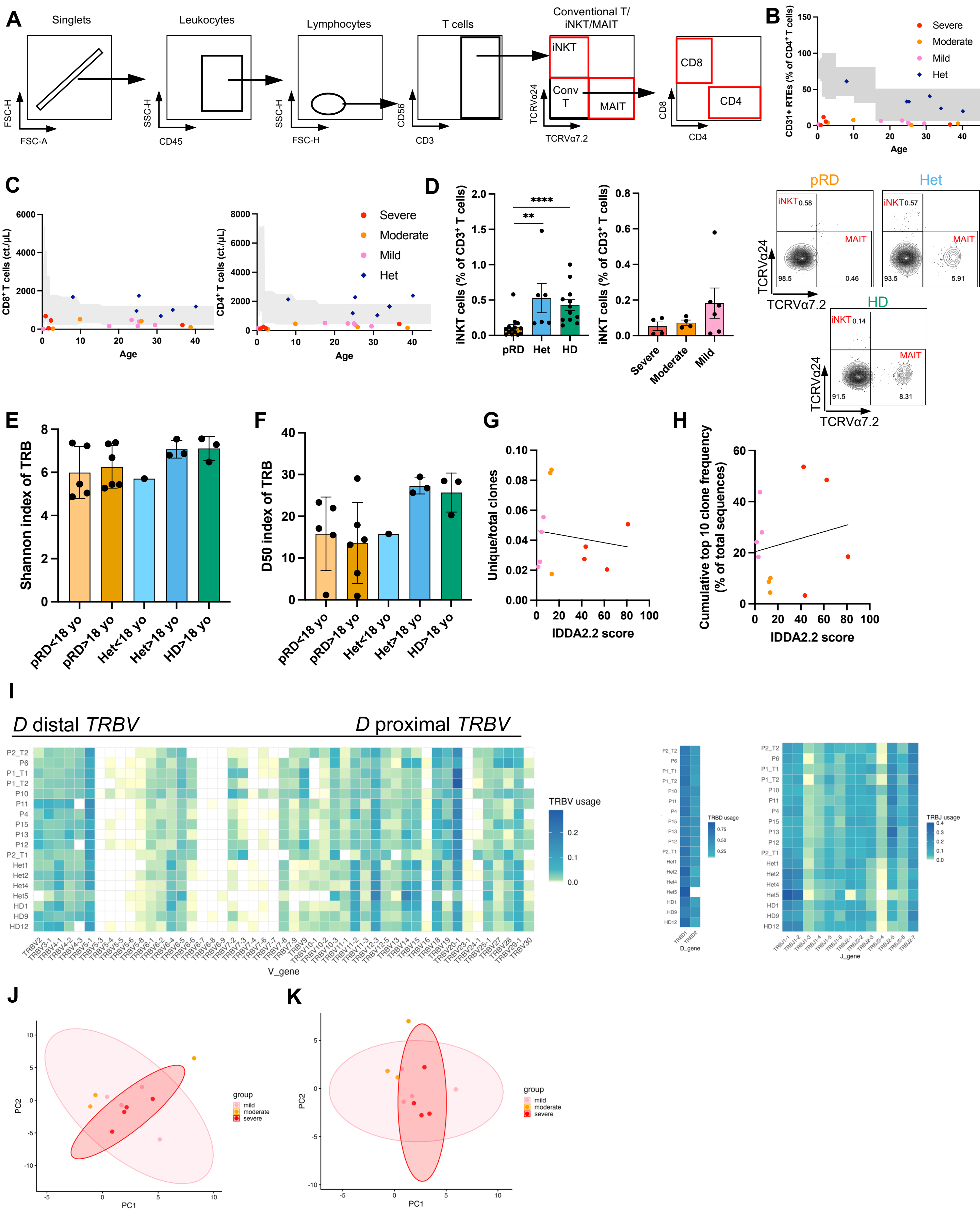


**Figure S3. Whole blood immunophenotyping for T cells and TCR repertoire characteristics in pRD. A)** Gating strategy for T cells. **B)** RTE in pRD patients (n=14) and RAG1 Hets (n=6) compared to the healthy range. **C)** CD8⁺ **(left panel)** and CD4⁺ **(right panel)** T cell count. **D)** Frequency of iNKT cells in pRD patients, RAG1 Hets and HDs (left panel) and in pRD patients grouped by disease categories: severe (n=4), moderate (n=4) and mild (n=6) **(right panel)** and their representative flow plots. **E)** Shannon index by age. **F)** D50 index by age. **G)** Correlation between frequency of unique clones and top 10 clones **(H)** and the IDDA2.2 scores. **I)** Heatmaps showing the frequency of unique TRBV **(left panel)** from distal to proximal to D region; TRBD **(middle panel)** and TRBJ gene segments **(right panel)**. **J)** PCA plot using unique TRBV gene segments of pRD patients. **K)** PCA plot for weighted TRBV gene segments of pRD patients.

Significance was calculated using Mann-Whitney tests; p≥ 0.05 was considered statistically significant. Correlation and p value was calculated by Spearman correlation and *p* test indicated by the ρ correlation coefficient.

Abbreviations: RTE: Recent Thymic Emigrant; iNKT: invariant Natural Killer T.

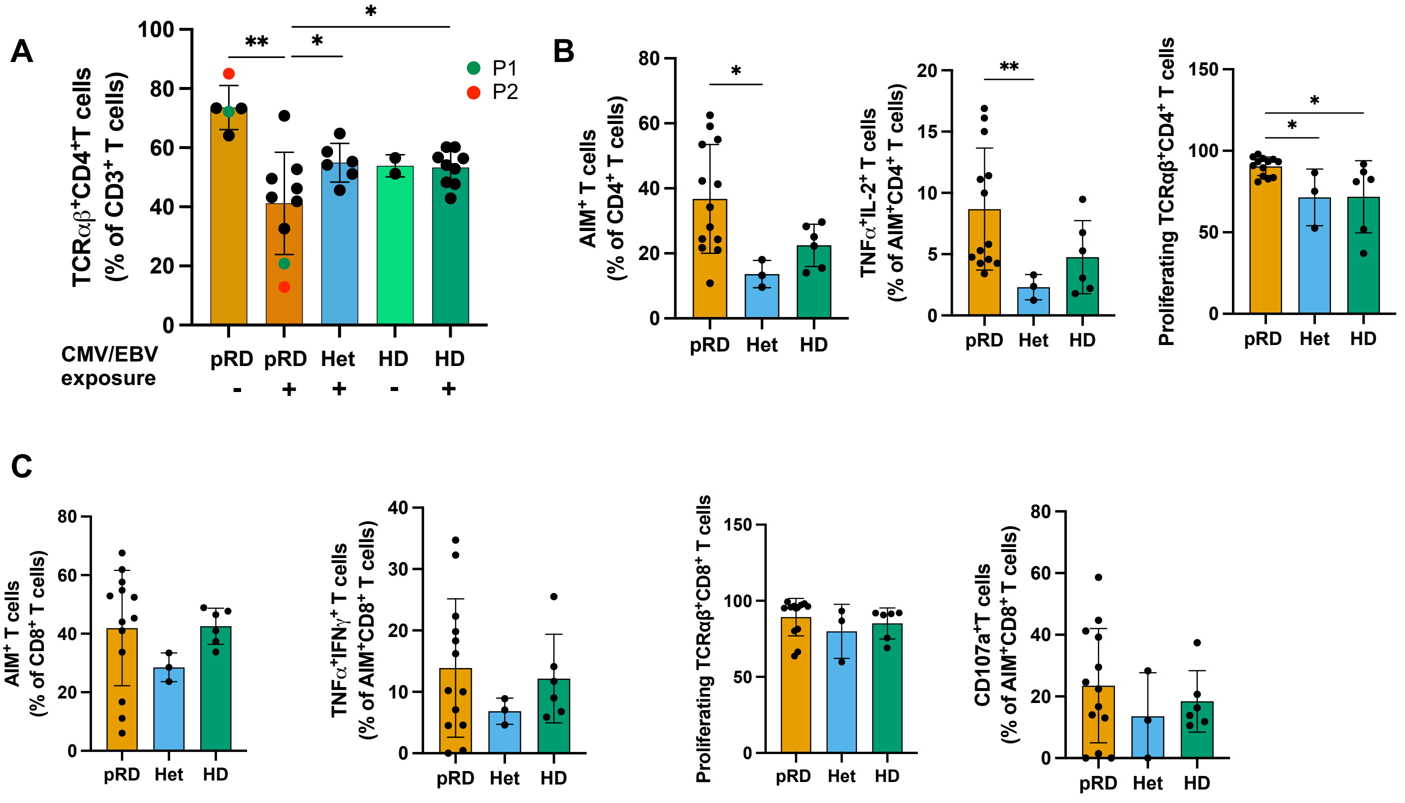


**Figure S4. CD4⁺ and CD8⁺ T cell responses to pan T cell activation. A)** TCRαβ⁺CD4⁺ frequency in whole blood of pRD timepoints (n=14), Hets (n=6) and HDs (n=12) segregated by Herpesvirus exposure. P1 and P2 timepoints comparing pre- and post-CMV disease are color-coded. **B)** AIM assay comparing pan stimulated CD4⁺ T cells **(left panel)**; cytokine secretion by pan peptide stimulated CD4⁺AIM⁺ T cells **(middle panel)**; proliferating capacity of CD4⁺AIM⁺ T cells upon pan T cell stimulation **(right panel)**. **C)** AIM assay comparing pan stimulated CD8⁺ T cells **(1st panel)**; cytokine secretion by pan stimulated CD8⁺AIM⁺ T cells **(2nd panel)**; proliferating capacity of CD8⁺AIM⁺ T cells upon pan T cell stimulation **(3rd panel)**; degranulation capacity measured by CD107a expression by CD8⁺AIM⁺ T cells upon pan T cell stimulation **(4th panel).**

Significance was calculated using Mann-Whitney tests; $p \geq 0.05$ was considered statistically significant.

Abbreviation: AIM: Activation Induced Marker.

**Supplementary Table 1:** Please contact authors for access.

| Panel | Antibody | Vendor |
|---|---|---|
| Panel A: Whole Blood immunophenotyping for B cells and myeloid cells | Anti-human-CD27, BV421 | BioLegend, Clone: M-T271, Cat# 356418 |
| | Anti-human-CD10, PE-Cy7 | BioLegend, Clone: HI10a, Cat# 312214 |
| | Anti-human-CD20, cFluor V547 | Cytek, Clone: 2H7, Cat# R7-20111 |
| | Anti-human IgM, PerCP-Cy5.5 | BioLegend, Clone: MHM-88, Cat# 314512 |
| | Anti-human CD38, BV711 | BioLegend, Clone: HIT2, Cat# 303528 |
| | Anti-human CD3, BV750 | BioLegend, Clone: SK7, Cat# 344846 |
| | Anti-human CD14, cFluor V450 | Cytek, Clone: M5E2, Cat# R7-20003 |
| | Anti-human-CD11c, PE | BioLegend, Clone: 3,9, Cat# 301606 |
| | Anti-human-CD56, BV570 | BioLegend, Clone: HCD56, Cat# 318330 |
| | Anti-human-CD24, BV650 | BD, Clone: ML5, Cat# 563720 |
| | Anti-human-CD19, cFluor R685 | Cytek, Clone: HIB19, Cat# R7-20117 |
| | Anti-human-HLA-DR, BV510 | BioLegend, Clone: L243, Cat# 307646 |
| | Anti-human-CD21, BV605 | BD, Clone: B-ly4, Cat# 740395 |
| | Anti-human-IgD, BV785 | BioLegend, Clone: IA6-2, Cat# 348242 |
| | Anti-human-CD16, cFluor R668 | Cytek, Clone: 3G8, Cat# R7-20069 |
| | Anti-human-CD123, cFluor R840/APC-Fire810 | BioLegend, Clone: 6H6, Cat# 306054 |
| | Anti-human-IgG, FITC | Cytek, Clone: M1310G05, Cat# 410720 |
| | Anti-human-CD45, cFluor R780/APC-Fire750 | BioLegend, Clone: 2D1, Cat# 306054 |
| Panel B: Whole Blood immunophenotyping for T cells | Anti-human-CCR4, BV605 | BioLegend, Clone: L291H4, Cat# 359418 |
| | Anti-human-CD45RA, cFluor V450 | Cytek, Clone: HI100, Cat# R7-20121 |
| | Anti-human-CCR7, BB515 | BD, Clone: 2-L1-A, Cat# 566764 |
| | Anti-human-CCR6, PE-TexasRed/PE-Dazzle594 | BioLegend, Clone: G034E3, Cat# 353430 |
| | Anti-human-TCR Va7.2, APC | BioLegend, Clone: 3C10, Cat# 351708 |
| | Anti-human-CXCR3, BV650 | BioLegend, Clone: G025H7, Cat# 353730 |
| | Anti-human-TCRgd, PE-Cy7 | BioLegend, Clone: B1, Cat# 331222 |
| | Anti-human-CD57, BV785 | BioLegend, Clone: QA17A04, Cat# 393330 |
| | Anti-human-CD28, APC-R700 | BD, Clone: CD28.2, Cat# 565181 |
| | Anti-human-CD3, BV750 | BioLegend, Clone: SK7, Cat# 344846 |
| | Anti-human-TCRab, PE-Cy5 | BioLegend, Clone: IP26, Cat# 306710 |
| | Anti-human-CXCR5, BV480 | BD, Clone: RF8B2, Cat# 566142 |
| | Anti-human-CD56, BV570 | BioLegend, Clone: HCD56, Cat# 318330 |
| | Anti-human-TCR Va24, PerCP-Cy5.5 | BioLegend, Clone: C15/TCR Va24, Cat# 360004 |
| | Anti-human-CD127, AF647 | BioLegend, Clone: A019D5, Cat# 351318 |
| | Anti-human-CD8, cFluor B532 | Cytek, Clone: SK1, Cat# R7-20123 |
| | Anti-human-CD4, cFluor B548 | Cytek, Clone: SK3, Cat# R7-20043 |
| | Anti-human-PD-1, BV421 | BioLegend, Clone: EH12.2H7, Cat# 329920 |
| | Anti-human-HLA-DR, BV510 | BioLegend, Clone: L243, Cat# 307646 |
| | Anti-human-CD31, BV711 | BioLegend, Clone: WM59, Cat# 303136 |
| | Anti-human-CD25, PE | BioLegend, Clone: BC96, Cat# 302606 |
| | Anti-human-CD27, APC-Fire810 | BioLegend, Clone: QA17A18, Cat# 393214 |
| | Anti-human-LAG3, RB705 | BD, Clone: T47-530, Cat# 570636 |
| | Anti-human-CD45, cFluor R780 | Cytek, Clone: 2D1, Cat# R7-20133 |
| Activation Induced Marker (AIM)/Intracellular Cytokine Secretion (ICS)/CellTrace Violet (CTV) assays | Live/Dead, e-fluor 780 | thermofischer/ebioscience, Clone: N/A, Cat# 65-0865-18) |
| | Anti-human-CD19, APC cy7 | Biolegend, Clone: HIB19, Cat# 302218) |
| | Ani-human-CD3, BV510 | Biolegend, Clone: SK7, Cat# 344827) |
| | Anti-human-CD4, PERCP5.5 | Biolegend, Clone: RPA-T4, Cat# 300530) |
| | Anti-human-CD8, BV421 | Biolegend, Clone: SK1, Cat#344748) |
| | Anti-human-CD8 (CellTrace assay), PE/Dazzle 594 | Clone: SK1, Cat# 344744) |
| | Anti-human-TCRgd, BV650 | Biolegend, Clone: B1, Cat# 331246) |
| | Anti-human-CD69, AF488 | Biolegend, Clone: FN50, Cat# 310916) |
| | Anti-human-CD137, PE/Cy7 | Biolegend, Clone: 4B4-1, Cat# 309818) |
| | Anti-human-CD134, PE/dazzle 594 | Biolegend, Clone: Ber-ACT35, Cat# 350020) |
| | Anti-human-IFNγ, PE | Biolegend, Clone: B27, Cat# 506507) |
| | Anti-human-CD107a (Lamp-1), AF647 | Biolegend, Clone: H4A3, Cat# 328612) |
| | Anti-human-TNFa, BV750 | Biolegend, Clone: MAb11, Cat# 502960) |
| | Anti-human-PD1, BV711 | Biolegend, Clone: EH12.2H7, Cat# 329928) |
| | Anti-human-CD57, BV785 | Biolegend, Clone: HNK-1, Cat# 393330) |
| | Anti-human-Tim-3, BV605 | Biolegend, Clone: F38-2E2, Cat# 345018) |
| | Anti-human-Ki67, AF647 | BD, Clone: B56, Cat# 561126) |
| | Anti-human-IL2, R718 | BD biosciences, Clone: MQ1-17H12, Cat# 567057) |
| | Anti-human-LAG3, BV480 | BD biosciences, Clone: T47-530, Cat# 746609) |
| | CellTrace Violet | thermofischer/ebioscience, Cat# C34557) |
| | Human FcX | Biolegend, Cat# 422302) |

**Table S2.** Antibodies used in whole blood immunophenotyping (Panel A, Panel B) and AIM/ICS/CTV assays.
